# The Urine Ammonium-pH Index is a Measure of Kidney Tubular Function

**DOI:** 10.64898/2026.08.06.26359870

**Authors:** Jesper Frank Andersen, Mads Vaarby Sørensen, Maria Chrysopoulou, Søren Gullaksen, Steffen Flindt Nielsen, Sandra Hummelgaard, Niklas Ayasse, Iben Skov Jensen, Louise Salomo, Natasja P. Simonsen, Jasmine CL Atay, Per Løgstrup Poulsen, Rikke Nørregaard, Liv Vernstrøm, Kathrin Weyer, Fatih Demir, Samuel Levi Svendsen, Alan M. Weinstein, Sebastian Nielsen, Marie Bodilsen Nielsen, Niels Henrik Buus, Henrik Birn, I. David Weiner, Markus Rinschen, Jens Leipziger, Peder Berg

## Abstract

Dysfunction of the tubulointerstitial compartment is a key driver of chronic kidney disease (CKD) progression. However, tubular function remains largely unaddressed by routine clinical assessment. Here, we show that the urine ammonium-pH index (uAPI), a composite of urinary ammonium and pH, reflects kidney tubular function and predicts kidney function decline.

Using acid/base, dietary, and potassium perturbations, segmental disruption of tubular ammonium handling, mathematical modelling, and data from patients with renal tubular acidosis, we identified defective ammoniagenesis as the main uAPI determinant. The uAPI was suppressed across four kidney disease models and dissociated from GFR. Kidney proteomics and single-nucleus RNA sequencing indicated downregulation of ammoniagenesis in proteinuric and diabetic kidney disease. In type 2 diabetes patients with preserved GFR, a reduced uAPI was associated with faster kidney function decline. In three CKD cohorts, low uAPI predicted CKD progression and significantly improved risk prediction.

Together, this positions the uAPI as a scalable, non-invasive measure of kidney tubular function.

## Introduction

Chronic kidney disease (CKD) is a complex and multifactorial disease affecting approximately 10% of the global population^1^ and is projected to become the fifth leading cause of years of life lost globally by 2040^2^. Early identification and management are critical as more therapeutic strategies become available^3^. Routine biomarkers, including estimated glomerular filtration rate (eGFR) and urinary albumin-creatinine ratio (uACR), predominantly assess glomerular function and filtration barrier integrity. In contrast, the tubulointerstitial compartment, which comprises most of the kidney parenchymal mass, remains largely unaddressed by routine clinical assessment^4^. The cells of this segment, especially in the proximal tubule, are among the most complex and energy-demanding throughout the entire body^5^. As such, they are known to be susceptible to damage through several different pathways^6^, e.g. deposition of albumin, fatty acids, or myoglobin^7–9^, or mitochondrial dysfunction due to increased glucose and sodium uptake^10^. Congruently, it is well established that tubulointerstitial damage and fibrosis in kidney biopsies are prognostic of subsequent kidney disease progression^11^. It has been suggested that the proximal tubule, and injury hereof, play a pivotal role in the development and progression of CKD^12,13^. Taken together, evaluating kidney health by glomerular functions alone might lead to erroneous conclusions regarding the actual state of the organ.

Ammonium generation via glutamine metabolization is a central and metabolically integrated function of the proximal tubule that produces *de novo* bicarbonate, which counteracts the daily load of non-volatile acids from diet and metabolism^14,15^. After production and secretion in the proximal tubule, ammonium is reabsorbed by the thick ascending limb, deposited into the medullary interstitium and further secreted into the collecting duct lumen and ultimately excreted with the urine^14,16^. Therefore, adequate ammonium excretion hinges on the integrated function of the entire tubular system, making urinary ammonium a possible measure of tubular health. Accordingly, low urinary ammonium excretion has previously been proposed as a marker of compromised acid handling^17^ and associated with increased risk of CKD progression^18,19^ and lower proximal tubule organic anion transport capacity^20^. However, as ammonium excretion is strongly modulated by the need for acid elimination^16^, low urinary ammonium excretion can either reflect diminished excretion capacity, a low demand for acid elimination, or a combination of these. This severely limits the use of ammonium as a measure of tubular function.

The urine ammonium-pH index (uAPI) was recently developed to address this limitation^21^. Using urinary pH to take the body’s need for acid excretion into account, the uAPI aims to address ammonium excretion capacity independent of the demand for acid elimination. An uAPI below the normal range indicates compromised kidney ammonium excretion and is independently associated with an elevated risk of CKD progression and kidney failure in CKD stages 3 and 4^21^. The main aim of this study was to determine the physiological and mechanistic factors governing the uAPI and to evaluate its clinical usability and risk prediction improvement beyond traditional markers of kidney function.

To examine the physiological determinants of the uAPI, healthy mice were subjected to controlled acid-base, potassium, and dietary challenges. Furthermore, mathematical modeling of ammoniagenesis along with a cohort of kidney transplant recipients were used. Next, the uAPIs sensitivity to segmental tubular dysfunction was investigated using four different knockout mouse models and reports of patients with renal tubular acidosis. Subsequently, the uAPI and its association with GFR were examined in five mechanistically distinct rodent models of kidney disease and in two human type 2 diabetes cohorts. Kidney proteomics and single-nucleus RNA sequencing complemented the functional data by detailing the molecular effect of proteinuric and diabetic kidney disease on the uAPI. Lastly, the clinical relevance and predictive value of the uAPI were addressed using three independent prospective CKD cohorts.

## Methods and materials

### Animals

All animal experiments were approved by the appropriate authorities (The Institutional Animal Care and Use Committee of the Gainesville Veterans Affairs Medical Center, and the University of Florida College of Medicine for studies in NBCe-1A/B, GS, and RhCg+RhBg mice and the animal experiments inspectorate, Denmark for the remaining). During housing, animals were kept in a 12-hour day/night cycle and had *ad libitum* access to drinking water and standard rodent chow unless otherwise stated.

#### C57Bl/6J and Balb/cByJ mice

Male and female C57BL/6J and Balb/cByJ mice were purchased from Janvier.

#### ZSF1 rats

Lean (ZSF1-Lepr^fa^/Crl) and obese male ZSF1 (ZSF1-Lepr^fa^ Lepr^cp^/Crl) rats were purchased from Charles River Laboratories and fed Purina No. 5008 diet (Research Diet, Brogaarden, Denmark).

#### Podocin mice

Compound heterozygous podocin^A286V/R231Q^ mice harboring loss-of-function mutations in the podocin gene, NPHS2, were used. The generation of this model was as previously described^22^. Podocin^A286V/R231Q^ mice were rederived at the Department of Biomedicine, Aarhus University, Denmark. Genetic background: C57BL/6NRj.

### NBCe1-A/B, proximal tubule-specific glutamine synthetase, collecting duct-specific RhBg + RhCg and NBCn1 KO mice

The generation of the mouse models was as previously described^23–26^. Genetic background: C57Bl/6J.

Table S1 provides an overview of the different animal models and experiments including species, strain, animal sex, methods for urine collection and NH^4+^ determination, and whether urinary pH and ammonium data were collected from previously published experiments.

### Randomization and blinding

Animals were randomly allocated to intervention groups where applicable, though no formal randomization algorithm was used. Investigators were not blinded to treatment or genotypes during animal experiments. Clinical data was collected blinded (for uAPI levels) from national registries and laboratory systems. Clinical samples were measured blinded (to clinical data and outcomes).

### Acid/base loading

Male C57Bl/6J mice were acid- or base-loaded by exposure to *ad libitum* drinking water containing 200 mM NH_4_Cl or 200 mM NaHCO_3_, respectively. Spot urine samples were collected under resting conditions and after 24 and 48 hours of acid-base loading.

NBCn1 mice were acid-loaded by exposure to 196 mM NH_4_Cl drinking water^26^.

NBCe1-A/B KO mice, GS KO mice, and RhBg + RhCg KO mice were acid-loaded by the addition of HCl to powdered rodent chow (0.4 M HCl, 1 ml/g chow), as described previously^23–25^.

### Dietary interventions

Male and female C57Bl/6J mice were fed either a standard rodent diet, a potassium-depleted diet (<0.02% K^+^, ssniff Spezialdiäten, Germany) for three days or a high-potassium diet (2% K^+^, made by adding KCl to a ssniff EF R/M control diet) for four days. Further, urine was collected from male and female C57Bl/6J mice fed before and after a simulated Western diet for 6 weeks (Research Diet, D19031901).

Nephrotic syndrome via doxorubicin injection and urine collection:

Male and female Balb/cByJ mice were injected with a single dose of doxorubicin (10 µg/g) administered via the tail vein. Experiments were terminated after 7 days.

### Angiotensin II infusion

Angiotensin II (750 ng/kg/min) was continuously infused via subcutaneous osmotic minipumps (Alzet 1004, Agnthos, Sweden) in C57Bl/6J mice of equal sex and age distribution for 4 weeks.

ZSF1 rats:

Obese rats were either treated with vehicle or empagliflozin for 15 weeks starting at 14-15 weeks of age. Empagliflozin did not ameliorate kidney disease progression^27^.

### Adenine-fed mice

Male and female C57Bl6/J mice of 10-12 weeks were fed either a 0.2% adenine, 6% casein diet (adenine diet, Brogaarden, Denmark) or a 6% casein diet (control diet, Brogaarden, Denmark) for up to three weeks.

### Patient cohorts

To assess the effect of kidney transplantation on the uAPI, the Aarhus University Hospital subset of CKD patients (n=126) undergoing renal transplantation from the multinational CONTEXT study^28^ was included.

To assess the association between dietary acid intake and the uAPI, we assessed the association between net endogenous acid production and the uAPI in the dietary intervention study, NNRD (n=60), conducted at the Department of Nephrology, Copenhagen University Hospital, Denmark^29^. The study randomized patients with CKD stages 3-4 to the “New Nordic Renal Diet” or a habitual diet for 6 months with 24-hour urine collections at baseline, after 2 weeks and monthly until end of study.

To assess whether the uAPI was affected in patients with type 2 diabetes mellitus and whether it was associated with kidney function decline, kidney cortical perfusion, kidney blood/plasma flow, and tubular damage markers, the uAPI was measured in patients with longitudinal data in two clinical studies, SiRENA (n=33 with diabetes, n=49 in total), conducted at the University Clinic in Hypertension and Nephrology, Gødstrup Hospital, Denmark^30^ and SEMPA (n=117, n=116 with longitudinal data), conducted at the Department of Endocrinology and Internal Medicine, Aarhus University Hospital, Denmark^31^. The SiRENA study included participants with type 2 diabetes mellitus, CKD or type 2 diabetes mellitus and CKD. The SEMPA study included patients with type 2 diabetes mellitus with cardiovascular disease, and/or heart failure, and/or CKD or high risk of cardiovascular disease. Inclusion and exclusion criteria for the studies are listed in the Supplemental Material.

For longitudinal analyses in CKD, patients with longitudinal data were included from three prospective clinical studies in which the uAPI was measured: RENVAS (n=79)^32^, PUMA (n=72)^33^, and SLEEP (n=75)^34^. These studies were conducted at the Department of Renal Medicine, Aarhus University Hospital, Denmark. RENVAS and PUMA included relatively unselected patients with CKD stages 3 and 4. SLEEP included patients with CKD stages 3-5, type 2 diabetes, and an uACR >30 mg/g not receiving kidney replacement therapy.

Inclusion and exclusion criteria for all studies are listed in the Supplemental Material.

The CONTEXT study (clinical trials registry number: NCT01395719) included participants from June 2011 to December 2014.

The NNRD study (clinical trials registry number: NCT04579315) included participants from November 2020 to November 2021.

The SiRENA study (EU Clinical Trials Register 2019-004303-12, 2019-004447-80 and 2019-004467-50) included participants from August 2021 to December 2022.

The SEMPA study (clinical trials registration information: EudraCT 2019-000781-38) included participants from August 2019 to February 2022.

RENVAS (clinical trials registry number: NCT01380717) included participants from March 2011 to March 2012.

PUMA included participants from April 2013 to December 2015.

SLEEP (clinical trials registry number: NCT04549324) included participants from September 2020 to January 2022.

### Longitudinal clinical data collection

Clinical data was collected from two type 2 diabetes mellitus (T2DM) cohorts (SEMPA and SiRENA) and three CKD cohorts (RENVAS, PUMA and SLEEP) databases, the local laboratory system, and the CROSS-TRACKS cohort. The latter being a population-based, open cohort containing routinely collected data from primary and secondary healthcare partners, combined with data from national registries. A full description of the cohort has been published previously^35^. Data on eGFR and time of initiation of long-term dialysis, kidney transplantation and death were collected from the time of urine sampling until the last follow-up (RENVAS: December 2018, PUMA: September 2023, SLEEP: March 2026, SiRENA: March 2026, SEMPA: March 2026). Follow-up was administratively censored at 5 years for SEMPA, SiRENA, and SLEEP. eGFR was calculated from plasma creatinine using the 2021 Chronic Kidney Disease Epidemiology Collaboration equation (CKD-EPI)^36^. The extent of missing data is shown in Table S2.

### Study approval

The seven included clinical studies were approved by the Committee on Biomedical Research Ethics for the Central Region of Denmark or Capital Region of Denmark. The studies were conducted in accordance with the ethical standards of the Danish National Research Committee and the World Medical Association Declaration of Helsinki. Written informed consent was obtained from all participants prior to any study procedures. Access to CROSS-TRACKS data was granted by the CROSS-TRACKS steering committee.

### Clinical outcomes

For longitudinal analyses in patients with type 2 diabetes (SiRENA and SEMPA cohorts) the following outcomes were assessed for patients with eGFR>60: annual eGFR decline, incident CKD (eGFR<60), and incident CKD with at least 25% reduction of eGFR. The risk for eGFR reductions of at least 40% and 50% were assessed in all patients with type 2 diabetes in the SiRENA and SEMPA cohorts.

For longitudinal analyses in the CKD cohorts, the main outcome was a composite CKD progression outcome defined as the first occurring event of either a ≥50% reduction in eGFR, eGFR<15 ml/min/1.73 m^2^, initiation of long-term dialysis, or kidney transplantation. Death was a censoring event. Secondary outcomes were as follows: 1) Kidney failure defined as the first occurring event of eGFR<15 ml/min/1.73 m^2^, initiation of long-term dialysis or kidney transplantation. Death was a censoring event. 2) CKD progression (≥50% reduction in eGFR, eGFR<15 ml/min, initiation of long-term dialysis, or kidney transplantation) or death.

### Urine collections

For rodents, urine was collected either as spot urine samples (C57Bl/6J, podocin, NBCn1, doxorubicin-treated mice, and adenine-treated mice), 24-hour metabolic cage collections under mineral oil (NBCe1-A/B, GS, RhBg + RhCg and NBCn1 mice), or 16-hour metabolic cage collections (ZSF1 rats). For human samples, urine was collected as spot urine samples (CONTEXT, SiRENA, SEMPA, and SLEEP) or 24-hour collections without mineral oil (RENVAS, PUMA, and NNRD).

### Patients with isolated proximal renal tubular acidosis, Fanconi syndrome and distal renal tubular acidosis

Urinary pH and ammonium data were collected from published reports of adult patients with proximal renal tubular acidosis^37,38^, Fanconi syndrome^39^, or distal renal tubular acidosis and their healthy controls^40–45^ where available. Patients were not included if they had decreased GFR (<60 ml/min/1.73 m^2^) or had mixed tubular defects, i.e. both proximal and distal defects in acid-base handling.

### Measurements

In C57Bl/6J mice, doxorubicin-treated mice, adenine-treated mice, podocin mice, ZSF1 rats and human cohorts, urinary ammonium and pH were measured using the same methods as previously described^46,47^. In short, urinary ammonium was measured with an Orion High-Performance Ammonia Ion-Selective Electrode (Thermo Scientific, Cat. No. 9512HPBNWP) with the use of an Ammonia pH-adjusting Ionic Strength Adjuster (Thermo Scientific, Cat. No. 951211). Urinary pH was measured using a micro pH electrode (pH-500, Unisense, Denmark) for mouse and rat samples and using a pH electrode (Metrohm, Herlev, Denmark) for human samples.

In NBCe1-A/B KO mice, GS KO mice, and RhBg/RhCg KO mice, urinary ammonium was measured using a commercially available kit (A7553, Pointe Scientific, Canton, MI) and urinary pH was measured with a micro pH electrode (ROSS semi-micro pH, Orion 8103BN)^23–25^.

In NBCn1 KO mice, urinary ammonium was measured with a fluorometric kit (MAK310, Sigma-Aldrich, USA). Urinary pH was measured using a micro pH electrode (pH-200, Unisense, Denmark)^26^.

The different methods for quantification of urinary ammonium have been shown to yield highly concordant results when directly compared^48,49^.

The uAPI was calculated from urinary ammonium concentration and pH as previously described^21^: uAPI = (log10(NH_4_^+^) x pH^3.6^)/40. The lower limit of NH_4_^+^ concentration was 1.1 mmol/L. For samples with NH_4_^+^ concentrations below 1.1 mmol/L, the concentration was assumed to be 1.1 mmol/L.

For patients with isolated proximal tubular acidosis, Fanconi syndrome, distal renal tubular acidosis and their controls, only urinary ammonium excretion values were available. Here, the uAPI was calculated from urinary ammonium excretion (mmol/day) and urinary pH as previously described^21^: uAPI = (log10(NH_4_^+^ excretion) x pH^2.7^)/10. Excretion based uAPI used NH_4_^+^ excretion in mmol/day in humans and in µmol/day in mice. Excretion-based uAPI was highly correlated with concentration-based uAPI (Pearson’s r=0.86 for CKD and 0.74 for controls, Figure S1)

For blood standard HCO_3_^-^ measurements in mice, blood was sampled from the tail vein during light isoflurane anesthesia with glass capillaries (CLINITUBES, Radiometer, Denmark) and immediately thereafter analyzed with an ABL90 Flex blood gas analyzer (Radiometer, Denmark).

In mice and rats, GFR was measured transcutaneously as previously described^50^. In short, the elimination of retro-orbitally injected fluorescein isothiocynate sinistrin (FITC-sinistrin; Fresenius Kabi, Graz, Austria) in conscious and freely moving animals over 60 min (mice) or 120 min (rats) was used as a measure of GFR.^51^

### Estimation of net endogenous acid production

Net endogenous acid production was calculated from estimated protein and potassium intake using a previously validated equation^52^: Net endogenous acid production (mmol/d) = −10.2 + 54.5 (protein intake [g/d] / potassium intake [mmol/d]). Protein intake was calculated from 24-hour urinary urea nitrogen excretion using the Maroni equation^53^ (protein intake = 6.25 x [urine urea nitrogen + 0.031 x body weight]). Potassium intake was estimated from 24-hour urinary potassium excretion.

### Mathematical modeling of whole rat kidney

Modeling of the uAPI during metabolic acidosis with varying degrees of proximal tubular ammoniagenesis was achieved using a previously developed model of whole rat kidney NH_4_^+^ fluxes^54^.

### Kidney proteomics and single-nucleus sequencing data

Proteome changes of prespecified proteins of interest (Table S3) in cortical and medullary kidney samples from podocin mice were reanalysed from Chrysopoulou et al^55^. Differential single-cell expression of prespecified genes of interest (Table S3) in human kidneys from controls and patients with diabetes was collected from the published Supplemental Material of Wilson et al.^56^

### Data exclusions

Urine samples contaminated by urease-producing bacteria become alkaline with abnormally high ammonium levels^57^ because of urea splitting which results in two NH_3_ per urea. NH_3_ will then form NH_4_^+^ resulting in a decrease in [H^+^] and an increase in [NH_4_^+^]. These samples were excluded as previously described^21^ (clinical samples: 1.1%, animal studies: 1.2%).

### Statistics

#### Software

GraphPad Prism 10.3.1 (GraphPad Software), Stata/BE 19 (StataCorp) and R 4.4.1 for Mac were used for statistical data analyses and graphical depiction of results. Proteomics data visualization was performed with Python (v3.9) using pandas (v2.1.4), NumPy (v1.24.2), matplotlib (v3.7.0), and seaborn (v0.11.2). In R, the packages riskRegression 2026.3.11, survival 3.8.6, rms 8.1.1, and nricens 1.6 were used to assess the incremental value of adding the uAPI to the KFRE. Pseudo-observation regression used prodlim 2026.3.11 and geepack 1.3.13. Figures were produced with ggplot2 4.0.2. Generative AI tools (Claude) were used to assist with code generation and refinement.

#### Data distribution

Data distribution was assessed by visual inspection of QQ plots. If the data could not be characterized as normally distributed and log-transformation could not adjust for this, non-parametric tests were used.

#### Sample size

The validation analysis in SLEEP was powered on the pooled adjusted hazard ratio of 7.15 reported previously^21^. Assuming a low uAPI prevalence of 67% and accounting for the correlation between the uAPI and the remaining model covariates (R² = 0.10), 14 primary outcome events were required for 90% power at a two-sided alpha of 0.05. Thirty-four events occurred, yielding 90% power to detect a hazard ratio of 3.49 or greater. For animal studies no formal power calculations were performed. It was assumed that a sample size of at least 6 per group would enable the identification of large physiologically relevant effects.

#### Statistical testing

When testing for statistical differences between two groups, unpaired or paired *t-tests* were used. Statistical differences between more than two groups were assessed using one-way ANOVA followed by correction for multiple comparisons. Comparisons between two groups with more than one intervention were evaluated using two-way ANOVA followed by correction for multiple comparisons. Regression analysis of two variables was performed using simple linear regression. Differential protein abundance between podocin^A286V/R231Q^ and control mice was assessed for each pre-specified protein using Welch’s t-test on log₂-transformed normalised intensities. Proteins with a Benjamini-Hochberg false discovery rate <0.05 (Welch’s t-tests, corrected within tissue across the pre-specified protein set) and |log₂ fold change| >0.2 were considered differentially expressed.

Changes in uAPI following kidney transplantation were assessed using a linear mixed-effects model with time as a fixed effect, participant as a random intercept and time as a random slope. Changes in uAPI and uACR as a function of age in podocin mice were assessed using a linear mixed-effects model with age (modeled as a 4-knot restricted cubic spline), genotype and the age-by-genotype as fixed effects, sex as a covariate, mouse ID as random intercept and age as random slope.

The association between a low uAPI and annual eGFR decline in patients with type II diabetes was assessed using a linear mixed-effects model with time, uAPI and time-by-uAPI interaction as fixed effects, participant as a random intercept and time as a random slope. Covariates were age, sex, BMI, log(uACR), and systolic blood pressure.

Cox proportional hazards models were constructed to assess the associations between baseline uAPI and incident CKD, incident CKD with at least 25% eGFR reduction, at least 40% eGFR reduction, at least 50% eGFR reduction, CKD progression, kidney failure and CKD progression or death. Cumulative incidence functions were used to graphically illustrate the risk of CKD progression stratified by uAPI (low vs. normal). The proportional hazards assumption was inspected by generating log-log plots. The associations between uAPI and outcomes were also assessed in competing risk models using Fine-Gray subdistribution hazard models, treating all-cause death as a competing risk. Models were constructed with and without adjustment for demographics (sex, age, and BMI), known risk factors for kidney function decline (uACR, eGFR, systolic blood pressure and diabetes status), and, in CKD cohorts, systemic acid-base status (total CO_2_). Covariates were selected based on theory and not according to how they altered the estimates or the models.

To assess the incremental value of the uAPI beyond the KFRE, the pooled RENVAS and PUMA cohorts served as the development set and SLEEP as an independent validation set. The 4-variable KFRE linear predictor was calculated using its published coefficients^58^:

Sum = (−0.2201 × ((age/10) − 7.036)) + (0.2467 × (sex − 0.5642)) − (0.5567 × ((eGFR/5) − 7.222)) + (0.4510 × (ln uACR [mg/g] − 5.137))

where sex was coded 1 for male and 0 for female. This linear predictor was entered as a fixed offset, so that no KFRE coefficient was re-estimated in any cohort. A reference model containing the offset alone was compared with an extended model additionally containing the uAPI as a continuous variable.

Baseline survivor functions were re-estimated in the development set by the Kalbfleisch–Prentice method, separately for each model and each conditional on its own linear predictor. Both baselines and the uAPI coefficient were then applied unchanged to SLEEP, so that no model parameter was estimated using SLEEP data. Absolute risk was computed as 1 - S₀(t)^exp(LP).

Hazard ratios and Harrell’s concordance index were estimated over the full follow-up. In SLEEP, all participants were administratively censored at 5.0 years, leaving no participants at risk at or beyond that horizon. Inverse-probability-of-censoring weights are therefore undefined at five years, whereas 39 of 75 remained at risk at four years. Measures requiring inverse-probability-of-censoring weighting at a fixed time (time-dependent AUC, Brier score, index of prediction accuracy, integrated discrimination improvement (IDI), and net reclassification index (NRI)) were accordingly evaluated at four years.

Discrimination was quantified by Harrell’s C and the time-dependent AUC, overall accuracy by the Brier score and the index of prediction accuracy (1 - Brier/Brier of a covariate-free Kaplan-Meier model), and reclassification by the IDI and the category-free NRI (Kaplan-Meier method for censored data). Between-model differences were obtained as paired contrasts with influence-function-based standard errors. Calibration was assessed by the calibration slope, regressing the outcome on the complementary log-log-transformed linear predictor and testing the coefficient against 1, and by the ratio of observed (Kaplan-Meier) to mean predicted risk. For the risk-gradient comparison, each model’s linear predictor was standardised within the cohort analysed and the hazard ratio per standard deviation obtained from a Cox model containing the standardised score as sole covariate. Confidence intervals for the IDI, NRI, and the ratio of risk gradients were obtained from 2,000 percentile bootstrap resamples, with model coefficients and baseline survivor functions held fixed throughout.

Cumulative incidence of CKD progression and absolute risk within strata were estimated with the Aalen-Johansen estimator, treating death without progression as a competing event, with pointwise 95% confidence intervals on the complementary log-log scale. To compare uAPI-stratified risk at similar levels of KFRE-predicted risk, participants from all three cohorts were pooled and stratified by tertile of KFRE-predicted risk and, within each tertile, by uAPI status (low, <13.4 a.u., versus normal). Risk differences between uAPI strata within tertiles were calculated as the difference between two independent Aalen-Johansen estimates.

Number needed to treat was calculated as the reciprocal of the product of the model-estimated Aalen-Johansen probability of CKD progression at five years and an assumed relative risk reduction of 30%

All tests were two-sided and performed at a significance level of 5%.

## Results

### The uAPI reflects the capacity for ammonium generation and excretion

For the uAPI to serve as a robust measure of tubular function, it must reflect the kidney’s capacity to excrete ammonium, rather than variations in acid excretory demand driven by diet or by other factors influencing renal acid-base handling.

#### The effect of systemic acid-base status on the uAPI

To directly assess the interplay between acid-base status and the uAPI, healthy C57Bl/6J mice were subjected to controlled acid or base loading.

At baseline, mice had an average uAPI of 29.3 a.u., which is ∼25% higher compared with healthy humans^21^, illustrating a right shift in the relationship between urinary ammonium and pH (Figure S2). Female mice had a ∼10% higher uAPI than male mice during resting conditions (Figure S3) consistent with higher ammoniagenesis in female mice^59^.

After 24 hours of acid loading via NH_4_Cl, urinary ammonium concentration increased and decreased urinary pH (Figure 1A). NaHCO_3_ loading induced the opposite changes (Figure 1A). Notably, the uAPI remained stable over a wide range of blood HCO_3_^-^ levels (10 to 30 mmol/L, Figure 1B). During 7 days of acid loading by dietary addition of HCl, the uAPI decreased slightly after 24 h in female mice but was otherwise stable throughout the period (Figure 1C). Similar stability was found using excretion-based uAPI (Figure S4)

**Figure 1:**
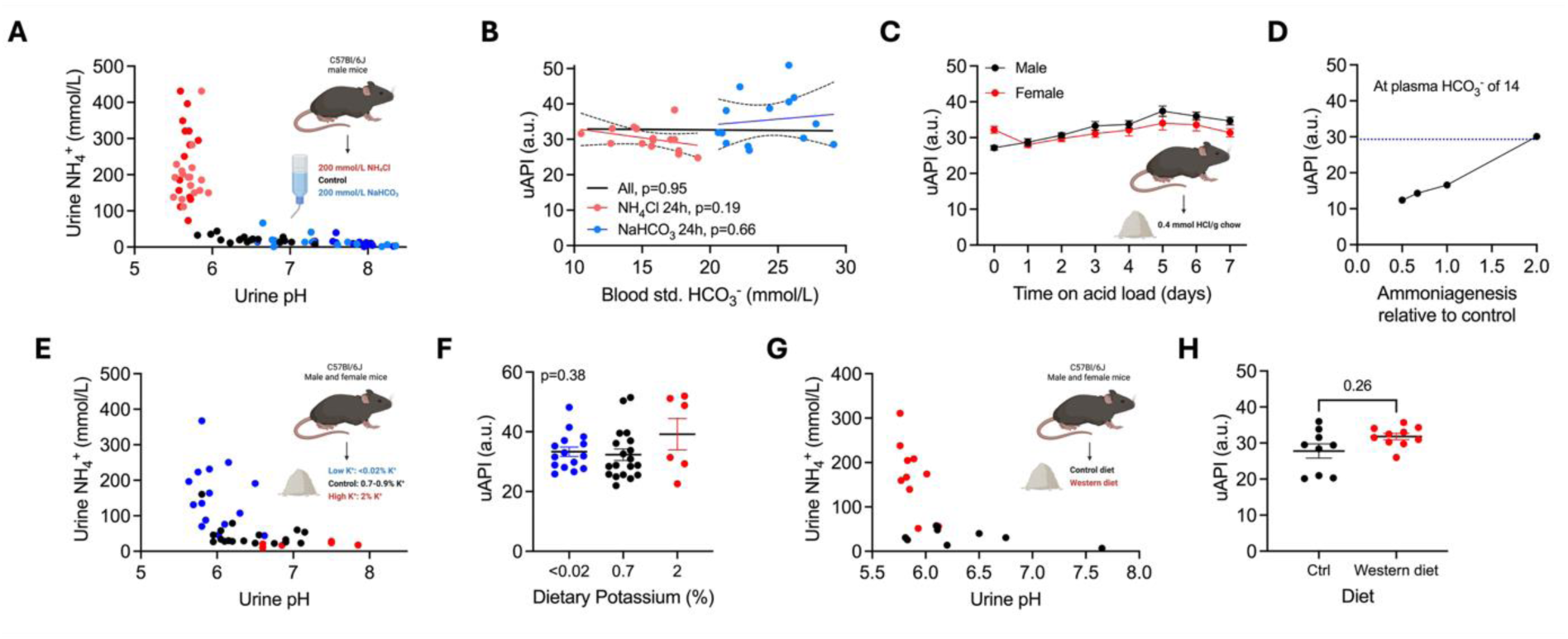
The uAPI is unaffected by acid-base, dietary, and potassium disturbances in healthy mice. (A) Urinary NH_4_^+^ as a function of urinary pH in mice exposed to control (black dots, n=17), NaHCO_3_-enriched (200 mmol/L, blue dots, n=15-17) or NH_4_Cl-enriched (200 mmol/L, red dots, n=15-17) drinking water for 24 and 48 hours. (B) uAPI as a function of blood standard HCO_3_^-^ (mmol/L) after 24 hours of acid (200 mmol/L NH_4_Cl drinking water, n=15) or base loading (200 mmol/L NaHCO3 drinking water, n=14). (C) uAPI in female (n=23-52) and male mice (n=20-47) before and during up to 7 days of dietary acid loading (0.4 mmol HCl/g chow). (D) uAPI as a function of ammoniagenesis relative to control in a mathematical model of whole rat kidney NH_4_^+^ and H^+^ flux. uAPI was calculated under systemic acidosis (plasma HCO_3_^-^ of 14 mmol/l). Unpublished calculations from^54^. (E-F): E) Urinary NH_4_^+^ as a function of urinary pH and F) uAPI in C57B6/J mice during control conditions (0.7-0.9% K^+^, n=19) or after dietary potassium depletion (3 days with a <0.02% K^+^ diet, n=15) or loading (4 days with a 2% K^+^ diet, n=6). (G-H): G) Urinary NH_4_^+^ as a function of urinary pH and H) uAPI in female and male mice during control conditions and after 6 weeks of being fed a Western diet (n=9-10, n=7 with paired observations). Statistical significances were assessed using simple linear regression within each group and for both groups with group as a covariate (panel B), Brown-Forsythe ANOVA test (panel F) and paired t-test (panel H).

Mathematical modeling of whole rat kidney ammonium and H^+^ flux provided further mechanistic insights. At a fixed degree of systemic acidosis (blood bicarbonate of 14 mmol/L), the uAPI was strongly determined by ammoniagenesis: a 50% reduction in ammoniagenesis severely suppressed the uAPI, while increased ammoniagenesis, the appropriate physiological response to acidosis, kept it within the normal range (Figure 1D).

Together, these data illustrate that, when able, the kidneys adequately adapt to an imposed acid or base challenge, leaving the uAPI largely unaffected.

#### Dietary effects on the uAPI

Generally, Western diets are low in, whereas plant-based diets are high in potassium and organic anions (base precursors). Both could affect acid-base and potassium balance and thus the uAPI.

In mice, potassium restriction increased and potassium loading decreased urinary ammonium as expected, but the uAPI remained unchanged (Figure 1E-F). Similarly, feeding healthy mice a Western diet did not alter the uAPI, despite large alterations in urine pH and ammonium (Figure 1G-H). In patients with CKD stages 3-4, the uAPI was stable across a wide range of estimated net endogenous acid production (Figure S5). These results indicate that the uAPI is largely independent of dietary patterns.

#### Association between plasma potassium and the uAPI

Plasma potassium levels modulate proximal tubular ammoniagenesis^60^. Hypokalemia stimulates, whereas hyperkalemia suppresses ammonium production. Lower plasma potassium was weakly associated with a higher uAPI in CKD stages 3-4 patients (p=0.02, r^2^=0.05, Figure S6), but not after adjustment for eGFR (p=0.65). When assessing the correlation within the normal range of plasma potassium, or using longitudinal repeated-measures data, no associations were found (p=0.20 and 0.24, respectively).

#### Effect of kidney transplantation on the uAPI

We previously found that in kidney transplant recipients, a low posttransplant uAPI associated with more tubular damage (more tubular VCAM1 expression in per-protocol day 6 biopsies) and poorer early graft function (higher risk of delayed graft function, longer time to halving of plasma creatinine, and lower day 5 mGFR) and worse late graft function (lower 12 month mGFR)^61^. Here, to determine whether a reduced uAPI primarily reflects impaired kidney function, we measured uAPI before and after kidney transplantation in CKD patients with preserved urine production.

Prior to transplantation, the uAPI was very low (median: 2.5 a.u.), driven by very low urinary ammonium levels. After transplantation, the uAPI increased rapidly and reached near-normal levels (>15 a.u.) within the first 24 hours and remained stable over 1 year of follow-up (Figure S7). Interestingly, delayed graft function and higher tubular VCAM1 expression were associated with much attenuated increases in the uAPI (Figure S7). While multiple parameters change post-transplantation, the rapid increase after graft implantation demonstrates that an improvement in kidney function acutely increases the uAPI.

### Targeted segmental dysfunction decreases the uAPI

To explicitly investigate whether impaired tubular function decreases the uAPI, mouse models with specific defects in ammoniagenesis or ammonium transport, but without other signs of kidney injury, were examined^23–26^. The uAPI was assessed under resting conditions and during increased demand for acid excretion, i.e. after 24 hours of acid loading.

#### Proximal tubule

NBCe-1 is a basolateral sodium bicarbonate cotransporter in the proximal tubule and KO severely impairs ammoniagenesis^24^. NBCe-1 KO mice had a strikingly lower uAPI under resting conditions compared with WT mice, a difference that became even more pronounced after acid loading (Figure 2B).

**Figure 2:**
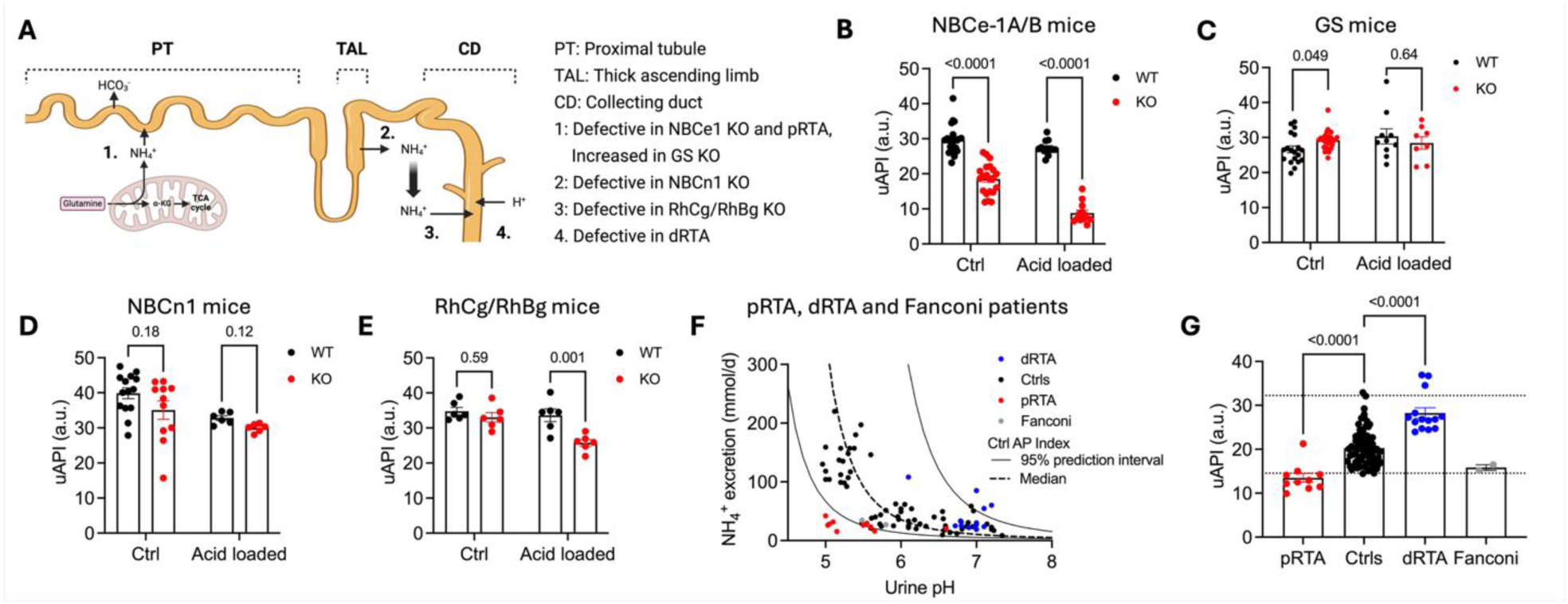
Segmental tubular dysfunction and proximal renal tubular acidosis decrease the uAPI. (A) Schematic overview of ammonium production and transport through the nephron. Sites of deficiencies corresponding to the disease models in panel B-F marked with numbers. (B-E) uAPI during control conditions and after 24 hours of acid-loading in B) NBCe-1A/B WT (n=12-21) and KO mice (n=12-21), C) GS WT (n=10-19) and KO mice (n=8-26), D) NBCn1 WT (n=6-14) and KO mice (n=6-11), and E) RhCg + RhBg WT (n=6) and KO mice (n=6). Mice were either dietary acid-loaded (0.4 mmol HCl/g chow, NBCe1, GS and RhCg + RhBg mice) or by NH_4_Cl-enriched drinking water (196 mmol/L, NBCn1 mice). (F) NH_4_^+^ excretion (mmol/day) as a function of urinary pH in healthy human controls (n=73) and patients with distal tubular acidosis (n=14), proximal tubular acidosis (n=10) and Fanconi syndrome (n=2). The dotted black line represents the association between urinary NH_4_^+^ and pH at the median uAPI of controls; the solid black lines represent the lower and upper limits of the 95 % prediction interval of healthy controls. (G) uAPI in the controls, dRTA, pRTA and Fanconi patients. In panel B-E, statistical differences were assessed by two-way ANOVA followed by Sidak’s multiple comparisons test. In panel G, differences were assessed using one-way ANOVA followed by Dunnett’s multiple comparisons test. uAPI values of GS mice were log10-transformed before statistical analysis. pRTA: proximal tubular acidosis. dRTA: distal tubular acidosis.

Glutamine synthetase (GS) KO mice lack the enzyme catalyzing the formation of glutamine from NH_4_^+^ and glutamate. This reaction decreases net ammonium production in the proximal tubule, and KO mice present with increased ammonium excretion during control conditions^25^. This was reflected in a slightly higher uAPI at baseline (Figure 2C).

#### Thick ascending limb

In the thick ascending limb ammonium is reabsorbed transcellularly, creating the interstitial corticomedullary ammonium gradient^26^. NBCn1 is a basolateral sodium bicarbonate cotransporter involved in maintaining the intracellular pH necessary for transepithelial ammonium transport from the lumen to the interstitium^26^. NBCn1 KO mice had tentatively lower, albeit not statistically significant, uAPI under baseline conditions (Figure 2D, overall genotype effect p=0.07). After acute NH_4_Cl loading by gavage, NBCn1 KO mice had a reduced uAPI (p=0.02, Figure S8). This aligns well with the phenotype of NBCn1 KO mice, which only experience a transient aggravation of metabolic acidosis when acid loaded^26^.

#### Collecting duct

Following interstitial accumulation in the thick ascending limb, ammonium is secreted in the collecting duct. Here, the Rhesus B and C gas channels play a pivotal role^23^. Therefore, the uAPI was investigated in mice with collecting duct-specific deletion of RhBg and RhCg. Under baseline conditions, no difference between KOs and WTs was observed, but after acid loading the uAPI decreased markedly in KOs while WTs remained stable (Figure 2E). This aligns well with the phenotype of collecting-duct specific RhBg + RhCg KO mice. They maintain normal acid-base balance during resting conditions but develop severe metabolic acidosis when acid loaded^23^.

Overall, similar findings were made using excretion-based uAPI calculations (the uAPI was strongly suppressed in NBCe-1A/B mice during control conditions and acid loading and in RhCg + RhBg KO mice during acid loading, Figure S9)

These data show that the uAPI decreases with dysfunctional NH_4_^+^ production and distal tubular handling. However, proximal tubular dysfunction suppresses the uAPI to a much greater extent compared with defective transport mechanisms in the distal tubular system.

#### Patients with tubulopathies

The uAPI was then investigated in published reports of patients with isolated proximal tubular acidosis (pRTA), distal tubular acidosis (dRTA) or Fanconi syndrome. Inherited isolated pRTA is most frequently caused by mutations in NBCe1. Therefore, it was hypothesized that these patients should present with a markedly reduced uAPI, despite normal GFR and other markers of kidney function. Conversely, patients with dRTA, whose disease is due to an acidification defect in the collecting duct, should present with a normal or even elevated uAPI due to the higher urinary pH.

The uAPI was markedly reduced in pRTA patients compared with controls (Figure 2F-G). Interestingly, two published cases of Fanconi syndrome also showed values below control range, despite normal creatinine clearance (Figure 2F-G). In contrast, dRTA patients had an elevated uAPI (Figure 2F-G).

The results recapitulate the dissociation between the uAPI and other markers of kidney function. Additionally, they illustrate the strong association between proximal tubular function and the uAPI and indicate its potential ability to differentiate between different etiologies of renal tubular acidosis.

### The uAPI decreases in kidney disease models independently of GFR

To investigate how different mechanisms of kidney injury affect the uAPI, multiple rodent models with either primarily glomerular, metabolic or tubular kidney disease were examined.

#### Glomerular dysfunction/damage

Compound heterozygous podocin mice (podocin^A286V/R231Q^) suffer from an extremely leaky filtration barrier and exhibit severe proteinuria and progressive kidney damage^22^. At 4 weeks of age, the uAPI was comparable to controls. However, at 6 weeks of age, the uAPI was significantly lower (Δ3.5 a.u., p=0.014) and decreased progressively as the mice aged (Δ21.2 a.u. vs. control at 19 weeks, p<0.0001) (Figure 3A). Interestingly, in control mice, the uAPI initially increased from 4 weeks of age until ∼10 weeks of age. This coincides with postnatal proximal tubule maturation which is thought to occur until at least week 8 in mice^62,63^. Furthermore, there was a remarkable temporal overlap between the decrease in the uAPI and the aggravation of proteinuria in podocin^A286V/R231Q^ mice (Figure S10).

**Figure 3:**
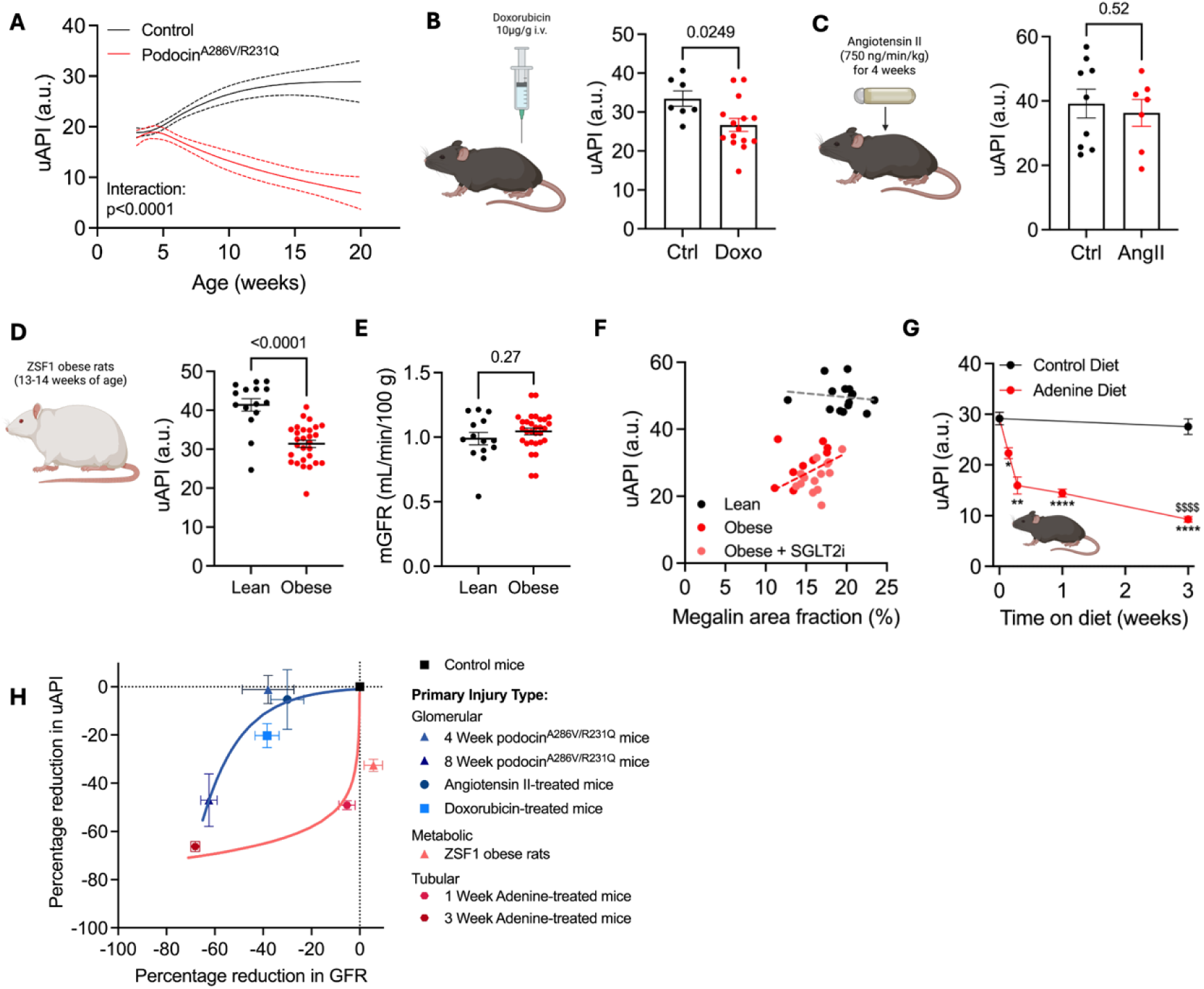
The uAPI is reduced in kidney disease and dissociates from GFR. (A) uAPI as a function of age in control mice (podocin^WT/R231Q^ and podocin^WT/A286V^ mice) and podocin^A286V/R231Q^ mice (n=92 measurements from 39 mice). (B) uAPI in saline (n=7) or doxorubicin injected (10 µg/g) Balb/cByJ mice (n=15) 7 days after injection. (C) uAPI before and after 4 weeks of angiotensin II infusion (750 ng/kg/min) via subcutaneous osmotic minipumps (n=7-9, n=6 with paired observations). (D, E) uAPI (D) and mGFR (E) in 13-14-week-old lean (n=14-15) and obese (n=28-30) ZSF1 rats. (F) uAPI as a function of megalin area fraction (%) in lean (n=15), obese (n=10) and obese SGLT2 inhibitor-treated (n=14) ZSF1 rats. (G) uAPI in mice before and after exposure to an adenine (0.2% adenine, 6% casein, n=7-8) or control diet (6% casein, n=5-8) for up to 3 weeks. (H) Percentage reduction in in uAPI as a function of percentage reduction in GFR in all rodent disease models. The relative reduction in GFR was estimated from the change in measured GFR in all models except doxorubicin treated mice, in which this was estimated from the increase in plasma creatinine. For podocin mice, previously published data on measured GFR was used^22^. Statistical differences were assessed using a linear mixed-effects model with age (modeled as a 4-knot restricted cubic spline), genotype and the age-by-genotype as fixed effects, sex as a covariate, mouse ID as random intercept and age as random slope (panel A), Student’s t-tests (panels B, D, E), a paired Student’s t-test (panel C), mixed-effects analysis followed by Dunnett’s multiple comparisons test (within-group comparisons, panel G) and two-way ANOVA followed by Sidak’s multiple comparisons test (between group comparisons, panel G), and linear regression analysis (panel F, for obese rats, treatment (+/- SGLT2 inhibitor treatment) was included as a covariate). *, **, ****: p<0.05, <0.01 and <0.0001, respectively, for baseline vs. adenine. $$$$: p<0.0001 for control vs. adenine at week 3.

The uAPI was also suppressed 7 days after induction of nephrotic syndrome by doxorubicin injection (Figure 3B). The model caused severe hypoalbuminemia, dyslipidemia, and tentatively increased plasma creatinine and blood urea nitrogen (Figure S11). No changes were seen after 4 weeks of angiotensin II infusion (Figure 3C) despite increased blood pressure, a decline in GFR, and increased albuminuria (Figure S12).

#### Metabolic dysfunction

In obese ZSF1 rats, a model of diabetic and hypertensive kidney disease^27^, the uAPI was markedly suppressed at 13-14 weeks of age (Figure 3D). Intriguingly, ZSF1 and lean control rats had comparable GFR at this timepoint (Figure 3E). In obese ZSF1 rats, a lower uAPI was associated with a smaller immunostaining positive area of the proximal tubular marker megalin at 29-30 weeks of age (Figure 3F, p=0.017, r^2^=0.35) along with higher urine excretion of the proximal tubular damage marker KIM-1 (Figure S13, p=0.006, r^2^=0.40). In contrast, no association was found for GFR or biopsy-evaluated fibronectin expression or glomerulosclerosis (Figure S13).

#### Tubular injury

In adenine-fed mice, a model that causes CKD by specifically damaging the tubulointerstitial compartment, the uAPI rapidly decreased (within days) compared with controls (Δ14.7 a.u. vs. baseline after 7 days, p<0.0001, Figure 3G).

Intriguingly, at early time points, uAPI reductions were much more pronounced relative to GFR reductions in metabolic and tubular injury models compared with glomerular injury models (Figure 3H). In the latter, decreases in the uAPI were mostly apparent only with severe reductions in GFR.

Overall, these data underline that several different mechanisms of kidney injury can diminish the uAPI. Importantly, the uAPI dissociates from GFR, decreasing either before or after GFR depending on the mechanism of injury (Figure 3H).

### Tubular dysfunction in human diabetic kidney disease

Next, the uAPI was investigated in two human T2DM cohorts (SiRENA and SEMPA, see Table S4 for characteristics).

Compared with healthy controls, patients with T2DM and preserved GFR had a markedly reduced uAPI (Figure 4A), despite comparable (even slightly higher) measured GFR and absence of albuminuria (Figure 4B-C). Patients with any CKD had a significantly lower uAPI compared with both controls and patients without CKD (Figure 4C).

**Figure 4:**
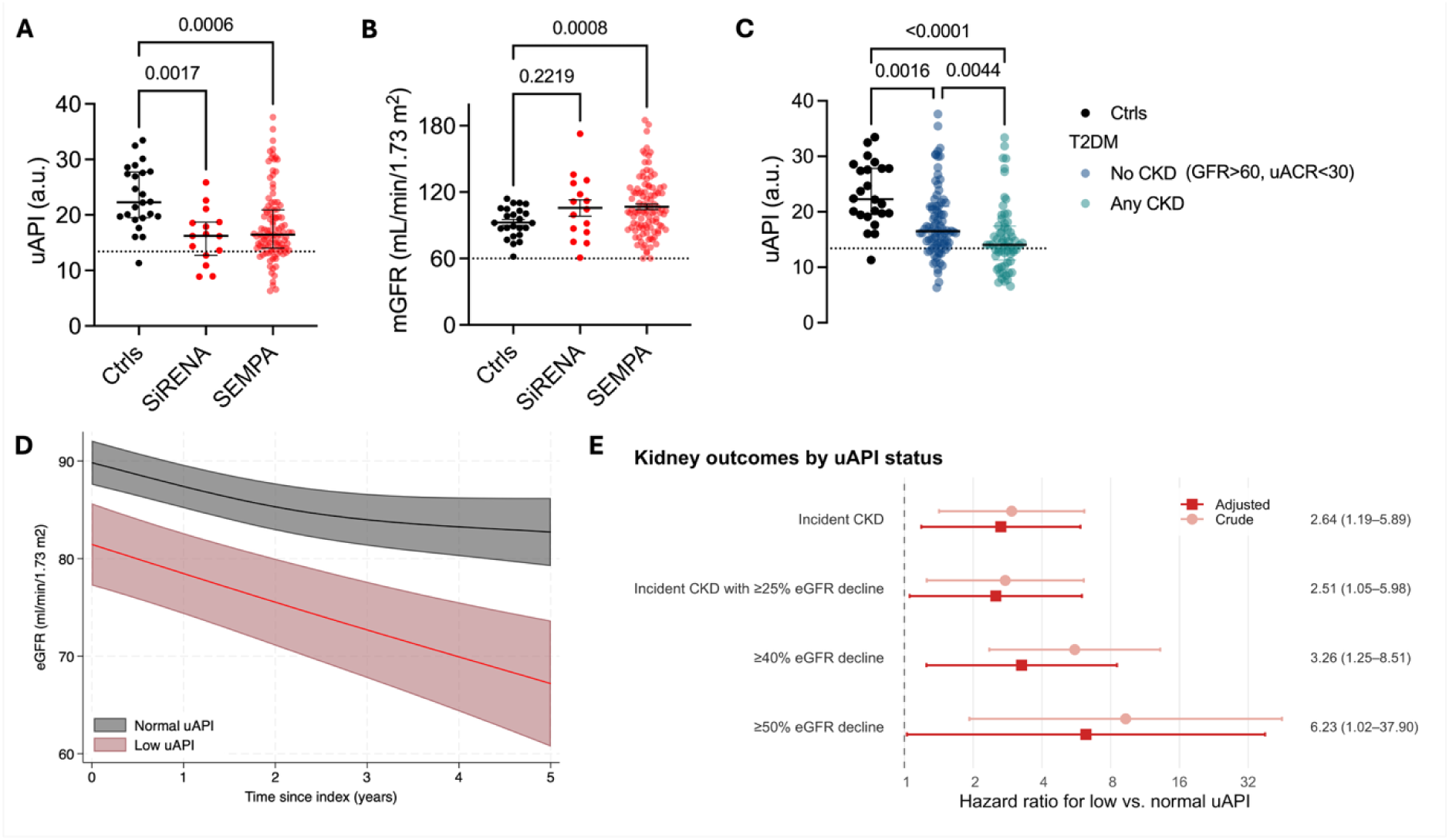
The uAPI is reduced in patients with type 2 diabetes and associates with higher risk of kidney outcomes. uAPI (A) and mGFR (B) in human controls (n=24-25) and T2DM patients with mGFR >60 mL/min/1.73 m^2^ from the SiRENA (n=15) and SEMPA (uAPI n=107, mGFR n=103) cohorts. (C) uAPI in the same cohorts stratified by no CKD (mGFR>60 and uACR<30, n=84) or any CKD (n=64). (D) eGFR (ml/min/1.73 m^2^) as a function of time (years) in T2DM patients with preserved eGFR at baseline in the same cohorts. (E) forest plot displaying adjusted hazard ratios for incident CKD, incident CKD with >25% eGFR decline, >40% eGFR decline and >50% eGFR decline for patients with low vs. normal uAPI in the same cohorts. Statistical differences were assessed using one-way ANOVA followed by Dunnett’s multiple comparisons test (panel A-C), a linear mixed-effects model with eGFR as outcome, time, uAPI, and time-by-uAPI interaction as fixed effects, time as a random slope, participants as a random intercept, an unstructured covariance matrix for within patient errors and age, sex, log(uACR), BMI and systolic blood pressure as covariates (panel D), and cox proportional hazard models adjusted for age, sex, BMI, eGFR, log(uACR), and systolic blood pressure (panel E). uAPI values in panel A+C were log transformed before statistical testing. Lines and error-bars represent median with IQR (panel A+C), mean with SEM (panel B), and mean with 95% CIs (panel D+E). The dotted line in panels A-C represents the previously established binary uAPI cut-off (<13.4 a.u.)^21^.

Intriguingly, the degree of uAPI reduction was not associated with renal blood flow, cortical perfusion, or tubular injury markers such as KIM-1 or NGAL (Figure S14).

Longitudinally, low uAPI at baseline (using our previously derived cut-off of 13.4 a.u.^21^) was associated with faster eGFR decline (adjusted difference for low vs. normal uAPI: 1.35 ml/min/1.73 m^2^/year, 95% CI: 0.13 to 2.57) and an increased risk of incident CKD (adjusted hazard ratio (aHR): 2.6, 95% CI: 1.2-5.9) and 40% eGFR decline (aHR: 3.3, 95% CI: 1.3-8.5) during follow-up (Figure 4D–E). Similarly adjusted subdistribution HRs were found in competing risk models with death as competing event (Figure S15)

### Multiple proteins/genes related to ammoniagenesis and H^+^/HCO_3_^-^ transport are decreased in podocin^A286V/R231Q^ mice and patients with T2DM

To investigate potential mechanisms underlying the suppressed uAPI in proteinuric kidney disease, the proteomic landscape of kidney cortex and medulla in 10-week-old podocin^A286V/R231Q^ and control mice was analyzed^55^. Specifically, differences in prespecified proteins known to be involved in ammoniagenesis or ammonium transport or H^+^/HCO_3_^-^handling along the tubular system were examined (Table S3^14,15^).

Many proteins related to proximal tubule ammoniagenesis and H^+^/HCO_3_^-^ handling were decreased in the podocin^A286V/R231Q^ mice (Figure 5). These included proteins involved in luminal (B^0^AT1, slc6a19) and mitochondrial glutamine import (Aralar1, slc25a12) and apical H^+^ secretion (V-type ATPase, atp6v0a4). At the basolateral side, NBCe1 (slc4a4), of which knockout severely decreased the uAPI, was downregulated. Metabolically, the cytosolic and mitochondrial malate dehydrogenases and key TCA enzymes^55^ were downregulated. No significant differences were found for thick ascending limb luminal import (NKCC2, slc12a1) or collecting duct basolateral import (RhCg) or apical secretion (RhCg) of ammonium/ammonia (Figure S16). However, key H^+^/HCO_3_^-^ transporting proteins were downregulated (e.g., the V-type H^+^-ATPase, pendrin, AE4, and the H^+^/K^+^ ATPase, Figure S16). Intriguingly, a publicly available single-nucleus RNA sequencing dataset of kidneys from people with and without T2DM^56^ allowed assessment of the genes encoding the proteins above. Here, SLC4A4 (NBCe1, proximal tubule cells), KCNJ1 and SLC4A7 (ROMK and NBCn1, thick ascending limb cells) and RhCg (collecting duct cells) were downregulated in T2DM (Figure S17-18). Furthermore, among >7000 annotated genes, SLC4A4 (NBCe1) was the 17^th^ most downregulated gene in injured proximal tubule cells. Interestingly, injured proximal tubule cells mirrored many of the findings in the podocin^A286V/R231Q^ mice (Figure S19).

**Figure 5:**
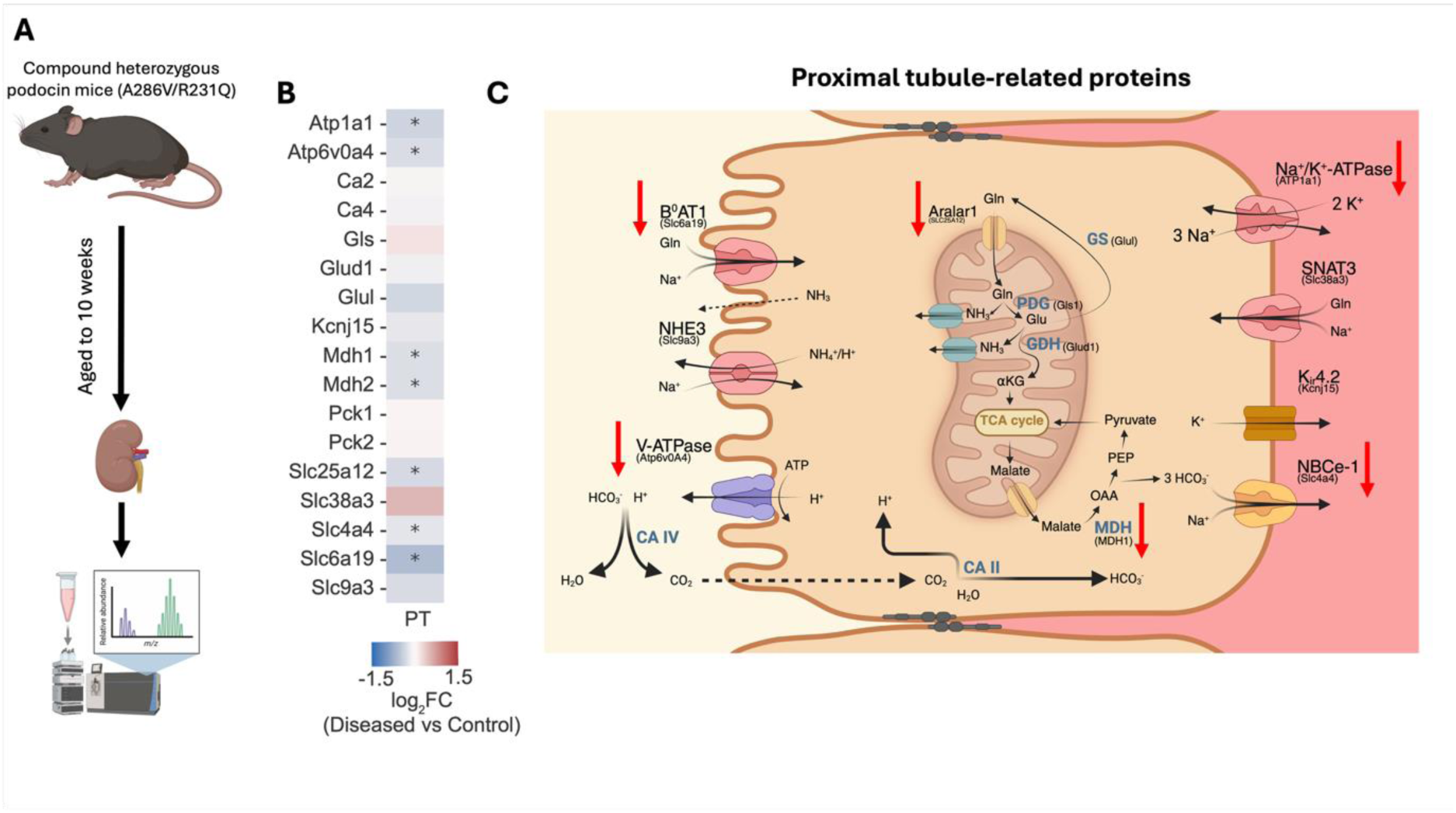
Numerous key proteins involved in proximal tubule ammoniagenesis and NH_4_^+^/H^+^/HCO_3_^-^ transport are downregulated in podocin^A286V/R231Q^ mice. (A) Cortical and medullary kidney tissue was harvested from 10-week-old podocin^A286V/R231Q^ (cortical n=4, medullary n=4) and podocin^R231Q/WT^ mice (cortical n=5, medullary n=6) and processed for proteomic analysis. (B) Differential cortical expression of a predefined (based on ^14,15^) selection of proteins central to ammoniagenesis, ammonium transport and HCO_3_^-^/H^+^ transport in proximal tubule cells. The color scale shows the log_2_ fold change in protein expression between the Podocin^R231Q/A286V^ and the Podocin^R231Q/WT^. Proteins with a Benjamini-Hochberg false discovery rate <0.05 (Welch’s t-tests, corrected within the pre-specified protein set) and |log₂ fold change| >0.2 are annotated with an asterisk. (C) Schematic representation of a proximal tubule cell with mitochondrial and cytosolic enzymes and transporters relevant to ammonium handling and luminal and basolateral transporters and channels relevant for ammonium and HCO_3_^-^/H^+^ transport. Red arrows indicate downregulation. PT: proximal tubule–related protein set.

Overall, tubular dysfunction, especially of the proximal tubule, appears to be a plausible explanation for the decreased uAPI in podocin^A286V/R231Q^ mice as well as in T2DM patients.

### Clinical utility of the uAPI in CKD

Having established the mechanistic basis of the uAPI, it was of priority to assess its clinical utility. This was done in three independent prospective CKD cohorts: RENVAS (n=79, CKD stages 3–4), PUMA (n=72, CKD stages 3–4), and SLEEP (n=75, CKD stages 3–5 with T2DM and uACR >30 mg/g). See Table S5 for characteristics. The association between a low uAPI and CKD progression was originally established in RENVAS and PUMA^21^. In this current study, we aimed to validate this finding in the independent SLEEP cohort and examine whether addition of the uAPI could improve risk prediction beyond the established Kidney Failure Risk Equation (KFRE)^58^.

In the SLEEP cohort, the cumulative incidence of CKD progression (defined as the first occurring event of a further 50% eGFR reduction, eGFR<15 or initiation of renal replacement therapy) was markedly higher in patients with a low uAPI (<13.4 a.u., Figure 6A) and a low uAPI was associated with an adjusted 5.7-fold higher hazard of CKD progression (Figure 6B). Large and significant estimates were also found for the secondary outcomes (i.e. kidney failure and CKD progression or death from any cause, Figure 6B). Consistent estimates were found using the uAPI as a continuous variable (Figure 6C), and across key subgroups, including sex (Figure S20-21). Similar adjusted subdistribution HRs were found in competing risk models with death as a competing event (Figure S22).

**Figure 6:**
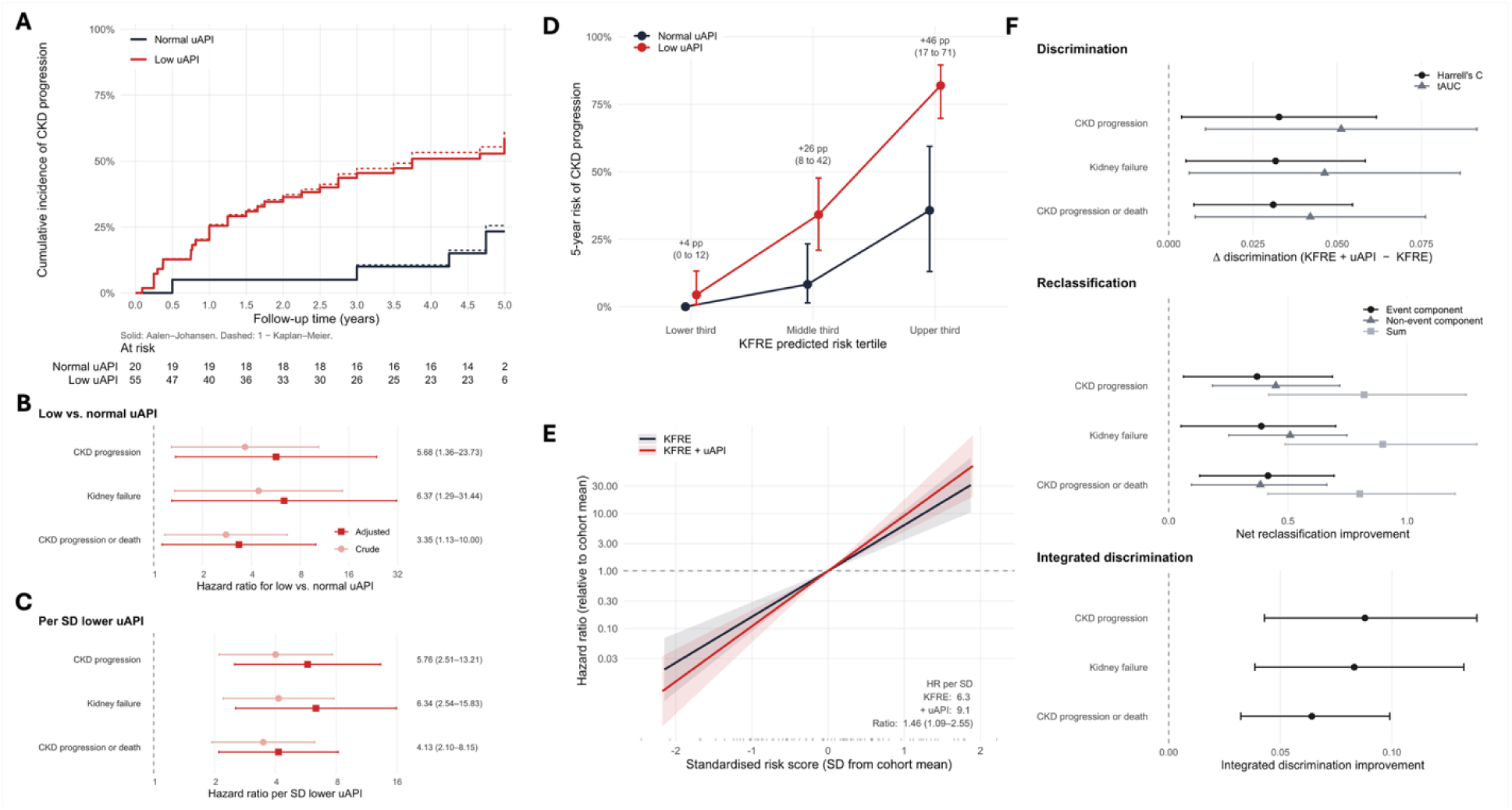
A low uAPI is strongly associated with CKD progression and improves risk prediction beyond the Kidney Failure Risk Equation. (A) Cumulative incidence of CKD progression (≥50% reduction in eGFR, eGFR<15 ml/min/1.73 m², initiation of long-term dialysis, or kidney transplantation) stratified for uAPI (low vs. normal, with low uAPI defined as <13.4 a.u.) in the SLEEP cohort (n=75). Solid lines represent the Aalen-Johansen estimator with death without progression as a competing event, dashed lines represent 1 - Kaplan-Meier with death as a censoring event. (B–C) Forest plots displaying crude and adjusted hazard ratios for CKD progression, kidney failure (eGFR<15 ml/min/1.73 m², initiation of long-term dialysis, or kidney transplantation), and CKD progression or death in the SLEEP cohort for B) low vs. normal uAPI and C) per standard deviation lower uAPI. (D) Five-year absolute risk of CKD progression within tertiles of KFRE-predicted risk, stratified for uAPI (low vs. normal), in patients from the RENVAS, PUMA, and SLEEP cohorts (n=226). Absolute risk differences between uAPI strata with 95% CIs are shown above each tertile. (E) Hazard ratio for CKD progression relative to the cohort mean as a function of the standardised risk score for the KFRE alone and the KFRE with the uAPI added, in the SLEEP cohort. Tick marks along the x-axis show the distribution of participants on the standardised KFRE + uAPI risk score. (F) Difference between the two models in discrimination (Harrell’s C and time-dependent AUC), net reclassification improvement (event component, non-event component, and their sum), and integrated discrimination improvement, in the SLEEP cohort. Hazard ratios were estimated using cox proportional hazards models and adjusted for age, sex, BMI, eGFR, log(uACR), blood total CO₂ and systolic blood pressure (panel B-C). Absolute risks were estimated using the Aalen-Johansen estimator (panel A, D). In panel E-F, the KFRE linear predictor was entered as a fixed offset, and the uAPI coefficient was estimated in the pooled RENVAS and PUMA cohorts and applied unchanged to SLEEP. Confidence intervals in panel E-F were obtained by percentile bootstrap (2,000 replicates). Lines and error-bars represent point estimates with 95% CIs.

To assess the potential value of using the uAPI, we assessed the incremental value of adding the uAPI to the Kidney Failure Risk Equation (KFRE)^58^. Before formally adding the uAPI, we assessed whether it separated risk among patients assigned similar risk by the KFRE. Within tertiles of KFRE-predicted risk, a low uAPI identified patients at substantially higher absolute risk of CKD progression (Figure 6D).

To quantify this formally, we fitted the KFRE using its published coefficients and added the uAPI as a continuous variable in RENVAS and PUMA. The resulting model, with both the KFRE coefficients and the uAPI coefficient frozen, was then applied unchanged to SLEEP and evaluated here.

In SLEEP, both models were adequately calibrated (Figure S23) and the addition of the uAPI resulted in a 45% steeper risk gradient (ratio 1.46, 1.09–2.55, Figure 6E). Across formal measures of predictive performance, addition of the uAPI improved discrimination in SLEEP (ΔHarrell’s C 0.033, 95% CI 0.004 to 0.062), with concordant improvements for kidney failure and for CKD progression or death and in measures of reclassification and overall accuracy (Figure 6F, Tables S6, and Figure S24). Changes in predicted risk for two hypothetical patients are shown in Table S7.

## Discussion

We recently published the development of the uAPI (initially termed the AB-score)^21^. Here, the mathematical derivation and conceptual background of the uAPI were detailed and it was found to associate with CKD progression in CKD stages 3-4. A low uAPI was interpreted as indicative of subclinical acid retention with detrimental effects on long-term kidney function. In this work, our main aim was to provide a firm physiological understanding of the uAPI, its underlying determinants, and its clinical utility.

Using a multilayered approach spanning controlled physiological experiments, a cohort of kidney transplant recipients, four genetic knockout models, patients with isolated tubular dysfunction, and five rodent disease models, supported by kidney proteomics and single-nucleus sequencing data, we establish the uAPI as a functional measure of kidney tubular function. Using longitudinal data from two T2DM cohorts and three CKD cohorts we assessed clinical utility.

Independent of systemic acid-base status, dietary patterns and potassium levels, the uAPI reflected the kidneys’ capacity for ammonium excretion, a task dependent on an orchestrated interplay between several compartments of the tubular system. Proximal tubular dysfunction, tubular injury, and diabetic kidney disease all severely diminished the uAPI independently of GFR. Congruently, when comparing different rodent disease models, the uAPI clearly dissociated from GFR decline (Figure 3H), suggesting that tubular functional impairment can precede the glomerular injury that decreased GFR and albuminuria eventually reflects. Therefore, a low uAPI in a patient with preserved or mildly reduced GFR may identify an individual on a trajectory to kidney failure who would not be flagged by current risk stratification, as illustrated in the T2DM cohorts.

Mechanistically, kidney proteomics and single-nucleus sequencing data supported tubular dysregulation of ammoniagenic pathways, as well as H^+^/HCO_3_^-^ transport, as key mechanisms underlying the suppressed uAPI in proteinuric and diabetic kidney disease. Finally, a low uAPI was associated with faster kidney function decline and CKD progression in T2DM patients with preserved kidney function and CKD patients.

In CKD, adding the uAPI to the established KFRE improved risk prediction in a cohort not used to develop the model. The gain in discrimination (ΔHarrell’s C 0.033) is comparable to that achieved by adding albuminuria to a model containing age, sex and eGFR alone (ΔHarrell’s C 0.018)^58^, and was accompanied by improvements in accuracy and reclassification. Further, concordance is computed over all comparable pairs and changes little when a marker reorders patients who are already close in predicted risk. The absolute risk differences within tertiles of KFRE-predicted risk (Figure 6D) show that the uAPI separates patients the KFRE ranks similarly. As an illustration, when applied to a treatment with a 30% relative risk reduction in kidney outcomes, such as an SGLT2 inhibitor, the predicted number-needed-to-treat over five years to prevent one progression event is approximately 3-4-fold lower in patients with a low uAPI as compared to a normal uAPI across KFRE-predicted risks of 5-50% (Supplementary Figure S25 and Table S8). These findings underscore the potential value of incorporating the uAPI into clinical assessment for risk stratification in CKD.

Several limitations of this study should be acknowledged. The clinical cohorts with longitudinal data are relatively small (n=149 and n=226 for T2DM and CKD, respectively) and were recruited only from Danish centers, limiting generalizability to other ethnic groups and healthcare settings. Most animal studies used both sexes but were not designed to specifically address sex differences. Data on the uAPI in pRTA and dRTA was collected from literature data and urine volume levels were not available in most instances to calculate concentration-based uAPI values. Finally, biopsy data for the CKD cohorts would have allowed for direct comparison between the uAPI and relevant histological patterns. Future studies with paired biopsy and uAPI data in patients with different CKD aetiologies would strengthen the molecular anchoring of the uAPI.

The physiological foundation of the uAPI also entails some important interpretive considerations. Large, acute changes in acid or base intake (e.g. bicarbonate supplementation or profuse vomiting) can promptly change urinary pH^64^, while changes in ammoniagenesis occur more gradually^14,23,24^. As a result, the uAPI could be misleading until appropriate proximal tubule compensation has occurred. Furthermore, certain pathologies, such as dRTA, that specifically impair distal tubular acidification increase the uAPI. Whether this could mask proximal tubular dysfunction or represents a genuine increased workload of the proximal tubule in dRTA, as observed in glutamine synthetase KO mice, remains to be determined. In addition, severe hypo- or hyperkalemia may also distort the uAPI.

Clinical implementation will require further external validation of the prognostic value in larger and more diverse patient cohorts. Future directions include evaluating the uAPI as a predictor of treatment response to kidney-protective medications such as SGLT2 inhibitors, GLP-1 receptor agonists or non-steroidal mineralocorticoid receptor antagonists. Additionally, the ability of the uAPI to predict acute kidney injury risk would be of significant interest.

In conclusion, the uAPI is a robust measure of tubular function. As a strong independent predictor of adverse kidney outcomes, the uAPI carries prognostic information that could aid in the earlier identification of high-risk T2DM patients, the separation of CKD patients with stable residual kidney function from those with a high risk of disease progression, and the enrichment of therapeutic trials to increase efficiency and reduce costs. We believe that inclusion of the uAPI offers a more comprehensive assessment of kidney function. As such, it could become a central measure for clinical decision-making.

## Acknowledgements

The work described in this article was supported by the Augustinus Foundation (2024-2508 to PB), the A.P. Møller Foundation for Medical Science (2024-01113 to PB), the NIH (R01-DK045788 and R01-DK107798 to IDW), the Department of Veterans Affairs (1I01BX000818 to IDW), the Novo Nordisk Foundation (NNF19OC0056043 and NNF24OC0095902 to MMR), the Aarhus University Research Fund (to MMR), and a Carlsberg Young Investigator Fellowship (to MMR). The funders had no role in study design, data collection and analysis, decision to publish, or preparation of the manuscript. The authors acknowledge the use of BioRender.com in creating schematics for Figures 1, 2, 3, and 5, Figure S7 and Figures S16-19. Adobe Illustrator (Adobe) was used for final Figure assembly of Figure 5B and Figure S16.

## Data availability

The source data and statistical source data (including exact p-values, etc.) underlying the Figures are provided with this paper. Complete individual-level clinical data used in this study cannot be made publicly available, as participant consent for data sharing was not obtained. Researchers who would like to inspect the raw data may apply for controlled access, subject to institutional approval. Data inquiries should be directed to the corresponding author.

