## Supplemental Material for "The Urine Ammonium-pH Index is a Measure of Kidney Tubular Function"

*[1]Department of Biomedicine, Aarhus University, Denmark, ,[2]Department of Endocrinology and Internal Medicine, Aarhus University Hospital, Denmark,[3]University Clinic of Hypertension and Nephrology, Gødstrup Hospital, Denmark, ,[4]Department of Renal Medicine, Aarhus University Hospital, Denmark, [5]Department of Cardio-Renal Pharmacology, Novo Nordisk, Måløv, Denmark, [6]Vth Department of Medicine, University Hospital Mannheim, Medical Faculty Mannheim of the University of Heidelberg, Mannheim, Germany, [7]Department of Molecular Medicine, University of Southern Denmark, Odense, Denmark, [8]Department of Nephrology, Herlev-Gentofte University Hospital, Denmark, [9]Department of Clinical Medicine, Aarhus University, Denmark [10]Department of Physiology and Biophysics, Weill Medical College of Cornell University, New York, United States, [11]Division of Nephrology Hypertension, and Renal Transplantation, University of Florida, Gainesville, FL USA, [12]Nephrology and Hypertension Section, Gainesville VA Medical Center, Gainesville, FL USA, [13]III. Department of Medicine, University Medical Center Hamburg-Eppendorf, Hamburg, Germany.*

*Corresponding author:

Dr. Peder Berg, Physiology, Department of Biomedicine, Health, Aarhus University, Denmark. **Address:** Høegh Guldbergs Gade 10, 8000 Aarhus C, Denmark, **mail:**

### Inclusion and exclusion criteria included clinical studies

#### CONTEXT

**Inclusion Criteria:**

- Age 18 and above
- Received information, signed consent
- Candidate for kidney transplantation from deceased donor

**Exclusion Criteria:**

- Can't give informed consent
- AV-fistula in the leg opposite the site where the graft will be placed
- Threatening ischemia in the leg
- If donor is a small child
- If the patient receives a double transplant

#### NNRD

**Inclusion Criteria:**

- Age >18
- Estimated glomerular filtration rate (eGFR) 20-45 ml/min
- Medically stable for two months prior to study start
- Written and verbally information is given
- Read, speak and understands Danish
- Written consent

**Exclusion Criteria:**

- Treatment with phosphate binders
- Metabolic disorders that requires specific dietary regulation
- Treatment with chemotherapy within the past 6 months
- Pregnancy and breastfeeding
- Food allergies
- Vegans

#### SiRENA

**Inclusion criteria**

**Study 1: Type 2 diabetes mellitus (DM2) and preserved kidney function**

- Aged 18 years or older
- Estimated glomerular filtration rate (eGFR) > 60 mL/min/1.73 $m^2$
- DM2 was diagnosed at least 1 year before inclusion and in stable medical antidiabetic treatment for at least 3 months
- Hemoglobin A1c (HbA1c) 48-70 mmol/mol (Diabetes Control and Complications Trial [DCCT] values 6.5%-8.6%)
- Fertile women were to use safe contraception

**Study 2: DM2 and chronic kidney disease (CKD)**

- Aged 18 years or older
- eGFR 20-60 mL/min/1.73 $m^2$

- DM2 was diagnosed at least 1 year before inclusion, and in stable medical antidiabetic treatment for at least 3 months
- HbA1c 48-70 mmol/mol. (DCCT values 6.5%-8.6%)
- Fertile women were to use safe contraception

**Study 3: Nondiabetic CKD**

- Aged 18 years or older
- eGFR 20-60 mL/min/1.73 $m^2$
- Fertile women were to use safe contraception

**Exclusion criteria**

**Study 1 and 2:**

- Type 1 diabetes
- Alcohol or substance abuse
- Pregnancy or breastfeeding
- Anamnestic or clinical signs of heart or liver failure
- Active cancers, aside from skin cancers (spinocellular or basocellular carcinomas)
- BMI > 35 kg/$m^2$
- Allergies or unacceptable side effects to the experimental treatment or background treatment
- If the investigator found the participant unfit to complete the trial.
- Previous kidney transplant
- Autosomal dominant polycystic kidney disease (ADPKD).

**Study 3: Nondiabetic CKD**

- Same as in study 1 and study 2
- DM2

#### SEMPA

**Inclusion criteria**

Individuals with type 2 diabetes with a HbA1c level of 48 mmol/mol (6.5%) or more and ability to give informed consent were eligible, fulfilling at least one criterion from one of the following categories:

50 years or older and at least one of the following:

- Prior myocardial infarction
- Prior stroke or transient ischemic attack
- Prior coronary, carotid, or peripheral arterial revascularization
- More than 50 % stenosis on angiography or imaging of coronary, carotid, or lower extremities arteries
- History of symptomatic coronary heart disease documented by e.g. positive exercise stress test or any cardiac imaging or unstable angina with electrocardiography (ECG) changes
- Chronic kidney impairment documented by estimated glomerular filtration rate below 60 ml/min per 1.73 $m^2$
- Chronic heart failure (New York Heart Association class II or III).

60 years or older and at least one of the following:

- Persistent microalbuminuria (30-299 mg/g) or proteinuria

- Hypertension and left ventricular hypertrophy by electrogram or imaging
- Persistent hypertension despite antihypertensive treatment
- Left ventricular systolic or diastolic dysfunction by imaging
- Smoking
- Ankle/brachial index less than 0.9

**Exclusion criteria**

- Estimated glomerular filtration rate < 45 ml/min per 1.73 $m^2$
- Treatment with a SGLT2-inhibitor, GLP-1-receptor agonist, or dipeptidyl-peptidase 4 inhibitor (DPP4-i) within 30 days before randomisation or insulin other than basal or 2 premixed within 30 days before randomisation. Participants are eligible after a 30-day wash-out period of SGLT-2i, GLP-1ra or DPP4-I treatment.
- A history of an acute coronary or cerebrovascular event within 90 days before randomisation.
- Planned revascularization of a coronary, carotid, or peripheral artery.
- Inability to give informed consent.
- Active cancer diagnosis other than basal cell carcinoma.
- Indication of liver disease (serum ALAT above 3 x upper limit).
- Bariatric surgery within the past two years and other gastrointestinal surgeries that induce chronic malabsorption.
- Treatment with systemic steroids at time of randomisation.
- Change in dosage of thyroid hormones within 6 weeks prior to screening.
- Alcohol or drug abuse within 3 months of informed consent that would interfere with trial participation or any ongoing condition leading to decreased compliance with study procedures or study drug intake.
- Chronic or acute pancreatitis
- Pregnancy or breastfeeding
- Allergy to either empagliflozin or semaglutide or any of the excipients contained in the drugs.

Changes to criteria for inclusion or exclusion were made as of September 2019 due to delayed recruitment and new data:

- Change of the inclusion criterion “HbA1c level 53 mmol/mol (7.0%)” to “48 mmol/mol (6.5 %)”
- Addition of the inclusion criterion “Chronic kidney impairment documented by estimated glomerular filtration rate below 60 ml/min per 1.73 $m^2$”
- Addition of the inclusion criterion “Persistent hypertension despite antihypertensive treatment“
- Change of the exclusion criterion “Estimated glomerular filtration rate < 60 ml/min per 1.73 $m^2$” to “Estimated glomerular filtration rate < 45 ml/min per 1.73 $m^2$”

#### RENVAS

**Inclusion criteria**

- age more than 18 years
- estimated glomerular filtration rate (eGFR) of 15–60 ml/min per 1.73 $m^2$, for at least 3 months,
- current use of antihypertensive medication or office BP at least 140/90 mmHg

- women in the fertile age had to use contraception.
- ability to give informed consent.

**Exclusion criteria**

- known allergies to study medications
- BP more than 130/80 mmHg determined by 24-h ambulatory measurements, despite treatment with maximum doses of four types of antihypertensive drugs
- nephrotic syndrome
- polycystic kidney disease
- known claustrophobia or MRI incompatible prostheses or pacemakers.

#### PUMA

**Inclusion criteria**

- age between 18 and 80 years
- newly referred to the renal outpatient clinic
- eGFR ranging from 15 to 60 ml/min/1.73m$^2$ for the last 3 months.
- ability to give informed consent.

**Exclusion criteria**

- acute kidney failure (defined as a decline in eGFR > 50% over the last 3 months)
- previous kidney transplantation
- polycystic kidney disease
- glomerulonephritis or renal vasculitis
- pre-existing cancer and/or life expectancy less than 18 months.

#### SLEEP

**Inclusion criteria**

- age above 18 years
- type 2 diabetes based on present antidiabetic treatment and an ICD diagnosis code of diabetes
- urine albumin-creatinine ratio (UACR) above 30 mg/g in at least 2 urine samples within the last 12 months
- estimated glomerular filtration rate (eGFR) 10–59 ml/min/1.73 m$^2$ (but not receiving renal replacement treatment) in at least 2 out of 3 separate measurements over a period of 12 months
- ability to give informed consent.

**Exclusion criteria**

- continuous positive airway pressure (CPAP) treatment within the last 3 months before recruitment
- previous coronary artery bypass grafting
- previous percutaneous coronary intervention (PCI).

### Supplementary Tables

#### Table S1: Animal Models

| Model | Species | Strain | Sex | Urine | $NH_4^+$ method | Previously published experiment |
|---|---|---|---|---|---|---|
| $NH_4Cl$, $NaHCO_3$ and control drinking water | Mouse | C57Bl6/J | Male | Spot | $NH_3$ Electrode | No |
| HCl chow | Mouse | C57Bl6/J | Male + female | 24h metabolic cages | Enzymatic | Yes[1-3] |
| Potassium loaded, restricted and control chow | Mouse | C57Bl6/J | Male + female | Spot | $NH_3$ Electrode | No |
| Western diet | Mouse | C57Bl6/J | Male + female | Spot | $NH_3$ Electrode | No |
| Podocin$^{A286V/R231Q}$ and control | Mouse | C57Bl6/J | Male + female | Spot | $NH_3$ Electrode | No |
| Doxorubicin injection | Mouse | Balb/cByJ | Male + female | Spot | $NH_3$ Electrode | No |
| Angiotensin II | Mouse | C57Bl6/J | Male + female | Spot | $NH_3$ Electrode | No |
| ZSF1 | Rat | ZSF1 | Male | 16h metabolic cages | $NH_3$ Electrode | Yes[4] |
| Adenine diet | Mouse | C57Bl/6J | Male + female | Spot | $NH_3$ Electrode | No |
| NBCe1 KO and WT | Mouse | C57Bl6/J | Male + female | 24h metabolic cages | Enzymatic | Yes[1] |
| GS KO and WT | Mouse | C57Bl6/J | Male | 24h metabolic cages | Enzymatic | Yes[3] |
| NBCn1 KO and WT | Mouse | C57Bl6/J | Male | Spot + 24h metabolic cages | Fluometric | Yes[5] |
| RhCg + RhBg KO and WT | Mouse | C57Bl6/J | Female | 24h metabolic cages | Enzymatic | Yes[2] |

#### Table S2: Extent of missing data

**Missing data**

| | SiRENA | SEMPA | SLEEP | RENVAS | PUMA |
|---|---|---|---|---|---|
| Age | 0% | 0% | 0% | 0% | 0% |
| Sex | 0% | 0% | 0% | 0% | 0% |
| BMI | 0% | 0% | 0% | 0% | 0% |
| Diabetes mellitus | 0% | 0% | 0% | 0% | 0% |
| Systolic BP | 0% | 0% | 0% | 0% | 0% |
| eGFR | 0% | 0% | 0% | 0% | 0% |
| Plasma $tCO_2$ | 48% | 100% | 0% | 0% | 0% |
| uACR | 0% | 0% | 0% | 0% | 0% |
| Urinary pH | 0% | 0% | 0% | 0% | 0% |
| Urinary $NH_4^+$ | 0% | 0% | 0% | 0% | 0% |
| uAPI | 0% | 0% | 0% | 0% | 0% |
| ACE inhibitor or ARB treatment | 0% | 0% | 0% | 1% | 0% |
| mGFR | 0% | 3.7% | NA | NA | NA |

Percentage of participants with missing data on key covariates at baseline. NA: Not applicable.

#### Table S3: Pre-specified proteins and genes of interest

| Proximal tubule-related | |
|---|---|
| **Gene name** | **Found?** |
| SLC6A19 | Yes |
| SLC38A3 | Yes |
| GLS1 | Yes |
| GLUD1 | Yes |
| MDH | Yes |
| PCK2 | Yes |
| PCK1 | Yes |
| GLUL | Yes |
| atp1a1 | Yes |
| SLC9A3 | Yes |
| SLC4A4 | Yes |
| AQP8 | No |
| KCNJ15 | Yes |
| CA2 | Yes |
| ATP6V0A4 | Yes |
| SLC25A12 | Yes |

| Thick ascending limb related | |
|---|---|
| **Gene name** | **Found?** |
| SLC12A1 | Yes |
| KCNJ1 | No |
| SLC4A7 | Yes |
| SLC9A4 | Yes |
| SLC9A1 | Yes |
| KCNJ10 | Yes |
| SLC9a3 | Yes |
| atp1a1 | Yes |

| Collecting duct related | |
|---|---|
| **Gene name** | **Found?** |
| Rhcg | Yes |
| Rhbg | Yes |
| ATP6V1B1 | Yes |
| ATP6V0A4 | Yes |
| ATP4A | No |
| Atp12a | Yes |
| ATP1A1 | Yes |
| CA2 | Yes |
| SLC4A1 | Yes |
| Slc26a7 | Yes |
| Slc12a2 | Yes |
| KCNJ10 | Yes |
| slc26a4 | Yes |
| slc4a9 | Yes |

#### Table S4: Characteristics of T2DM cohorts for longitudinal analyses

| Cohort | SiRENA | SEMPA |
|---|---|---|
| Participants | 33 | 116 |
| Age, years (IQR) | 72 (67-74) | 70 (65-74) |
| Female, n (%) | 6 (18) | 22 (19) |
| BMI, kg/m$^2$ (SD) | 30 (4) | 31 (5) |
| Diabetes mellitus, n (%) | 33 (100) | 116 (100) |
| Systolic BP, mmHg (SD) | 141 (13) | 141 (19) |
| eGFR, mL/min/1.73m$^2$ (SD) | 69 (26) | 90 (14) |
| Measured GFR, mL/min/1.73m$^2$ (SD) | 65 (29) | 102 (29) |
| Venous $tCO_2$, mmol/L (SD) | 26 (2) | NA |
| UACR, mg/g (IQR) | 77 (16-446) | 15 (7-66) |
| Urine pH, pH units (SD) | 5.9 (0.8) | 5.8 (0.7) |
| Urine $NH_4^+$, mmol/L (IQR) | 7.7 (6.1-11.4) | 22 (12.5-30.5) |
| Urine uAPI, a.u. (IQR) | 13.2 (9.9-16.2) | 16.3 (13.8-20.9) |
| ACE inhibitor or ARB treatment, % | 73% | 83% |
| Follow-up time, years (IQR)[a] | 3.5 (3.3-3.8) | 5 (5-5) |
| Events, incident CKD, n (%)[b] | 2 (13) | 28 (26) |
| Event rate, incident CKD, per 1000 patient years (95% CI)[b] | 37 (9-146) | 63 (43-91) |
| Events, incident CKD with ≥25%, n (%)[b] | 2 (13) | 24 (23) |
| Event rate, incident CKD with ≥25%, per 1000 patient years (95% CI)[b] | 37 (9-146) | 52 (35-78) |
| Events, ≥40% eGFR decline, n (%) | 7 (21) | 16 (14) |
| Event rate, ≥40% eGFR decline, per 1000 patient years (95% CI) | 66 (31-138) | 30 (18-49) |
| Events, ≥50% eGFR decline, n (%) | 2 (6) | 7 (6) |
| Event rate, ≥50% eGFR decline, per 1000 patient years (95% CI) | 18 (4-70) | 13 (6-27) |

[a]Follow-up time was defined as the total time a patient was followed, i.e. until loss-to-follow-up or death. [b]Incident eGFR<60 was only evaluated in patients with a baseline eGFR above or equal to 60 (n=15 in SiRENA and n=107 in SEMPA). One participant (SiRENA: 0, SEMPA: 1) had a measured uAPI but no follow-up after the index visit and could contribute no time at risk and was excluded from this table and from all longitudinal analyses.

#### Table S5: Characteristics of CKD cohorts for longitudinal analyses

| Cohort | RENVAS | PUMA | SLEEP |
|---|---|---|---|
| Participants | 79 | 72 | 75 |
| Age, years (IQR) | 65 (53-71) | 71 (66-75) | 74 (67-80) |
| Female, n (%) | 21 (27) | 27 (37) | 22 (29) |
| BMI, kg/m$^2$ (SD) | 27 (4) | 29 (5) | 31 (5) |
| Diabetes mellitus, n (%) | 19 (24) | 30 (42) | 75 (100) |
| Systolic BP, mmHg (SD) | 130 (15) | 140 (24) | 141 (16) |
| eGFR, mL/min/1.73m$^2$ (SD) | 37 (13) | 40 (13) | 34 (13) |
| Venous t$CO_2$, mmol/L (SD) | 28 (4) | 25 (4) | 26 (3) |
| UACR, mg/g (IQR) | 62 (0-689 | 53 (9-448) | 590 (192-1754) |
| Urine pH, pH units (SD) | 5.9 (0.6) | 5.9 (0.6) | 5.7 (0.6) |
| Urine $NH_4^+$, mmol/L (IQR) | 7.2 (4.8-9.5) | 7.3 (4.9-9.8) | 8.9 (4.6-15) |
| Urine uAPI, a.u. (SD) | 12.7 (5.3) | 13 (5.9) | 11.7 (4.3) |
| ACE inhibitor or ARB treatment, % | 87% | 65% | 82% |
| Follow-up time, years (IQR)[a] | 7.3 (6.8-7.6) | 8.5 (5.1-9.4) | 4.8 (4.6-5) |
| Events, CKD progression, n (%) | 29 (37) | 22 (31) | 34 (45) |
| Event rate, CKD progression, per 1000 patient years (95% CI) | 76 (53-109) | 57 (37-86) | 140 (100-196) |
| Events, kidney failure, n (%) | 28 (35) | 17 (24) | 31 (41) |
| Event rate, kidney failure, per 1000 patient years (95% CI) | 72 (50-104) | 43 (26-68) | 125 (88-178) |
| Events, CKD progression or death, n (%) | 39 (49) | 39 (54) | 40 (53) |
| Event rate, CKD progression or death, per 1000 patient years (95% CI) | 102 (74-139) | 98 (72-134) | 165 (121-224) |

[a]Follow-up time was defined as the total time a patient was followed, i.e. until loss-to-follow-up or death. Three participants (RENVAS: 2, PUMA: 1, SLEEP: 0) had a measured uAPI but no follow-up after the index visit and could contribute no time at risk. Four participants had contaminated samples (RENVAS: 0, PUMA: 2, SLEEP: 2). all are excluded from this table and from all longitudinal analyses.

#### Table S6: Incremental value of adding the uAPI to the KFRE

| **Cohort** | **RENVAS + PUMA** | | |
|---|---|---|---|
| **Model** | **KFRE** | **KFRE + uAPI** | **Difference** |
| **Harrell's C** | 0.837 | 0.857 | 0.020 (-0.002 to 0.042) |
| **tAUC** | 0.894 | 0.921 | 0.027 (0.005 to 0.048) |
| **Brier score** | 0.11 | 0.10 | -0.01 (-0.024 to 0.003) |
| **Index of predictive accuracy** | 0.45 | 0.50 | 0.05 (-0.02 to 0.11) |
| **Cohort** | **SLEEP** | | |
| **Model** | **KFRE** | **KFRE + uAPI** | **Difference** |
| **Harrell's C** | 0.841 | 0.874 | 0.033 (0.004 to 0.062) |
| **tAUC** | 0.856 | 0.908 | 0.051 (0.011 to 0.091) |
| **Brier score** | 0.15 | 0.11 | -0.033 (-0.056 to -0.010) |
| **Index of predictive accuracy** | 0.37 | 0.51 | 0.14 (0.04 to 0.23) |

Discrimination and accuracy metrics for prediction of CKD progression of the kidney failure risk equation (KFRE) applied to the RENVAS + PUMA cohort before and after addition of the continuous uAPI. Both models were then, without refitting, applied to the independent SLEEP cohort. Harrell's C utilized full follow-up while time-dependent AUC (tAUC), Brier score and index of predictive accuracy were evaluated at year 5 in RENVAS + PUMA and year 4 in SLEEP.

#### Table S7: Examples of predicted risk using the KFRE and the KFRE + uAPI

| Patient | 1 | 2 |
|---|---|---|
| Age, years | 70 | 70 |
| Sex | Male | Male |
| eGFR, ml/min/1.73 $m^2$ | 35 | 35 |
| uACR, mg/g | 300 | 300 |
| uAPI, a.u. | 20 | 8 |
| Five-year risk KFRE-predicted | 28.8% | 28.8% |
| Five-year risk KFRE + uAPI-predicted | 8.8% | 39.8% |

Two hypothetical patients identical on every KFRE variable, showing predicted five-year risk with and without the uAPI, using the frozen model developed in RENVAS and PUMA and the continuous uAPI.

#### Table S8: Number needed to treat by KFRE-predicted risk tertile and uAPI status.

| KFRE tertile | uAPI | n | Events | 5-year risk | 95% CI | NNT |
|---|---|---|---|---|---|---|
| Lower | Normal | 30 | 0 | 0% | not estimable | - |
| Lower | Low | 46 | 2 | 4.4% | 30–153 | 75.6 |
| Middle | Normal | 27 | 2 | 8.3% | 16–90 | 40.3 |
| Middle | Low | 48 | 16 | 34.1% | 6.9–16 | 9.8 |
| Upper | Normal | 14 | 5 | 35.7% | 5.2–23 | 9.1 |
| Upper | Low | 61 | 50 | 82.0% | 3.7–4.6 | 4.1 |

Five-year risk of CKD progression estimated by the Aalen-Johansen estimator (as in Figure 6D), with death without progression treated as a competing event, in participants pooled across RENVAS, PUMA and SLEEP (n=226). Tertiles were defined on KFRE-predicted five-year risk; a low uAPI was defined as below 13.4 a.u. Number needed to treat is the reciprocal of the product of the five-year risk and the assumed relative risk reduction, and is here shown for a relative risk reduction of 30%. Confidence intervals are percentile bootstrap (2,000 resamples of the cohort, model held fixed). No events occurred before five years among participants with a normal uAPI in the lowest tertile, so the number needed to treat is not estimable in that stratum. NNT, number needed to treat.

### Supplementary Figures

#### Figure S1: Comparison of excretion- and concentration-based uAPI

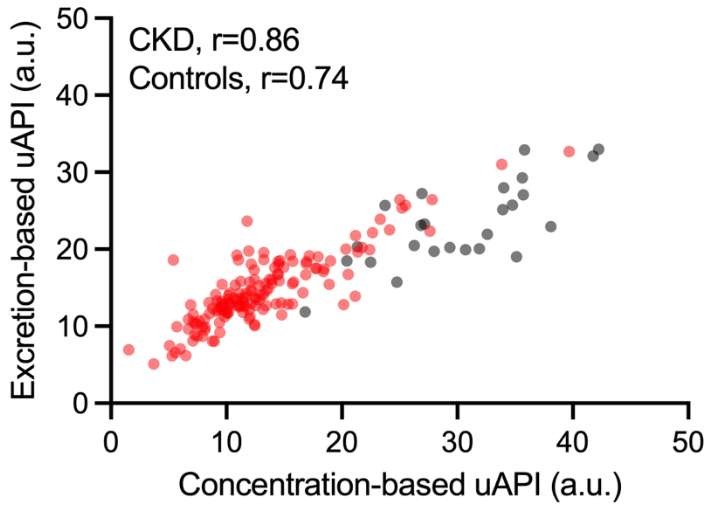

Comparison between concentration-based uAPI (uAPI=(log10([NH4+]) x $pH^{3.6}$)/40) and excretion-based uAPI (uAPI=(log10(NH4+ excretion) x $pH^{2.7}$)/10). Correlation was assessed using Pearsons r.

#### Figure S2: Comparison of the uAPI in Mice and Humans

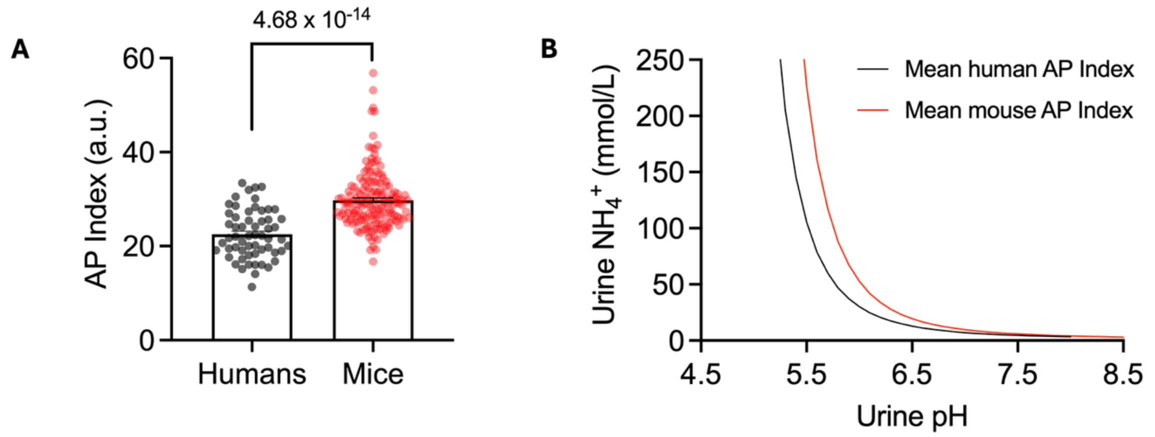

A) Comparison of uAPI in mice and humans. Statistical difference was assessed using students t-test. B) association between urine $NH_4^+$ and urine pH at the mean uAPI of humans (black) and mice (red).

#### Figure S3 Comparison of the uAPI in Male and Female Mice

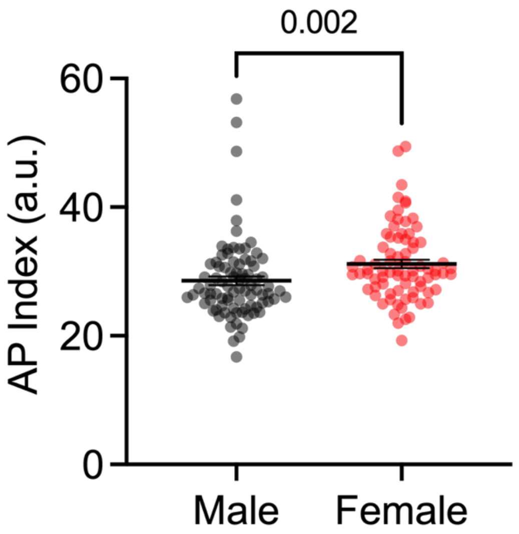

Comparison of uAPI between male (black, n=86) and female mice (red, n=78). Statistical difference was assessed using a t-test. Data was log-transformed before statistical assessment.

#### Figure S4: Excretion-based uAPI during acid-loading in mice

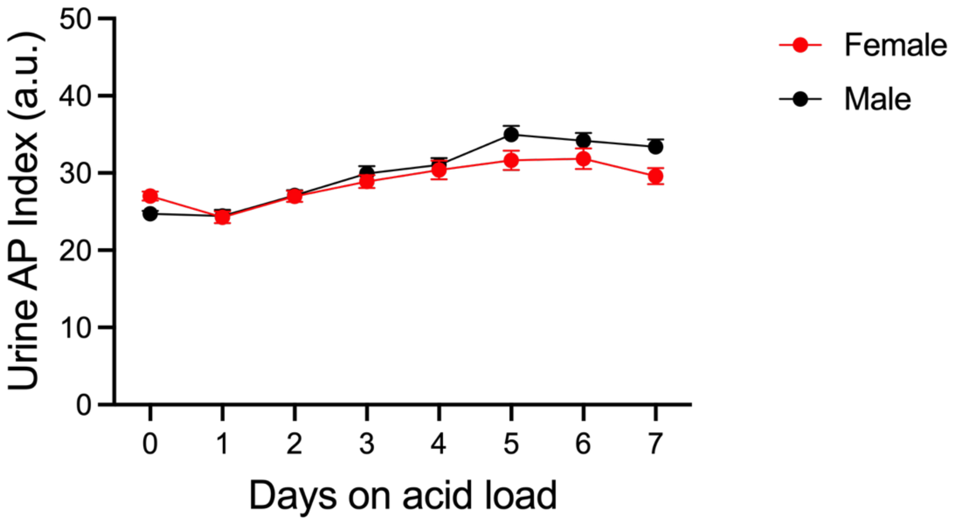

Excretion-based uAPI in female (n=23-52) and male mice (n=20-47) before and during up to 7 days of dietary acid-loading (0.4 mmol HCl/g chow).

#### Figure S5: Association between net endogenous acid production and the uAPI

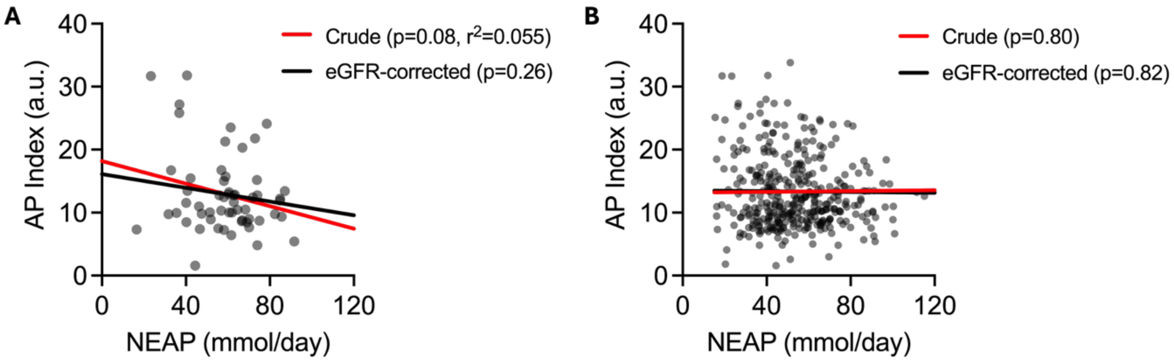

Association between net endogenous acid production and the uAPI in the NNRD study at baseline (A, n=58) and at all visits (B, n=366 observations from 60 patients). Associations were evaluated using simple linear regression (A) or a mixed-effects model for repeated measures with net endogenous acid production as fixed effect, patient ID as random intercept and visit as a random slope without (red line) or with (black line) eGFR as a co-variate (B).

#### Figure S6: Association between the uAPI and plasma potassium in CKD

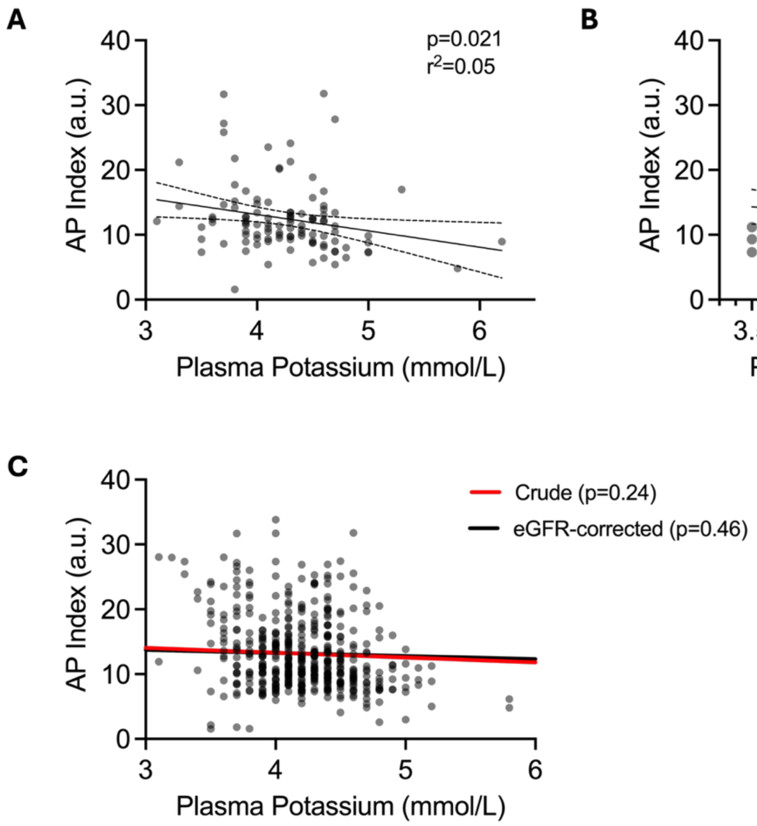

A-B) Association between plasma potassium and the uAPI in CKD stage 3-4 patients from the NNRD and PUMA study at baseline across all available measurements (A, n=107) and within the reference interval of plasma potassium (B, n=76). C) Association between plasma potassium and the uAPI in CKD stage 3-4 patients from the NNRD study at all visits across all available measurements (n=389 observations from 59 patients). Associations were evaluated using simple linear regression (A+B) or a mixed-effects model for repeated measures with plasma potassium as fixed effect, patient ID as random intercept and visit as a random slope without (red line) or with (black line) eGFR as a co-variate (C).

#### Figure S7: Changes in the uAPI after kidney transplantation

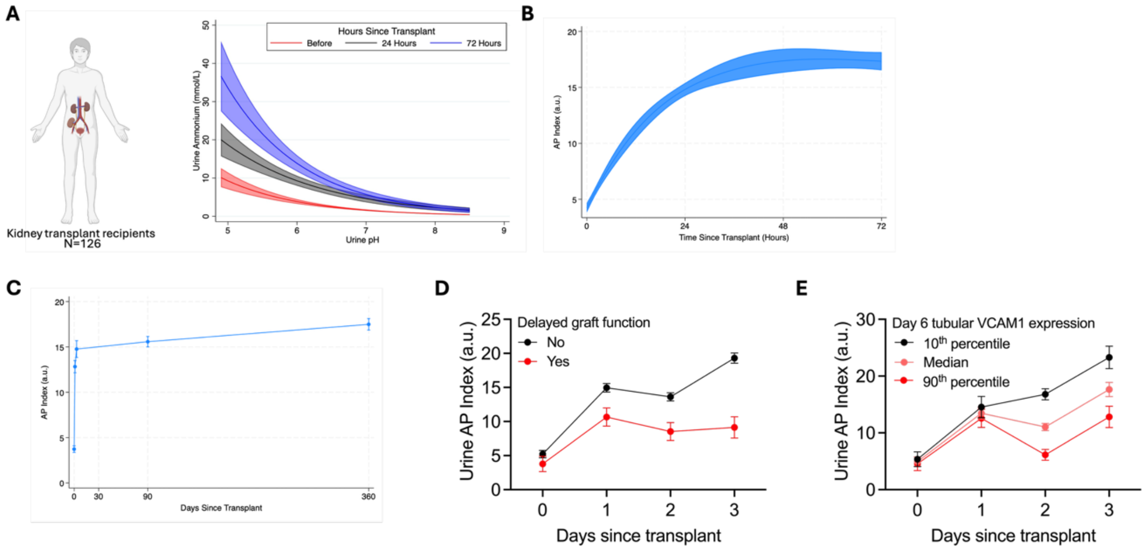

A) association between urine ammonium and urine pH before, 24, and 72 hours after kidney transplantation. B) time-dependent changes in the uAPI following a kidney transplant during the first 72 hours. C) time-dependent changes in the uAPI following a kidney transplant during the first year posttransplant. D) time-dependent changes in the uAPI following a kidney transplant during the first 72 hours stratified for the occurrence of delayed graft function (defined as need for dialysis within the first 7 days). E) time-dependent changes in the uAPI following a kidney transplant during the first 72 hours stratified for day 6 tubular VCAM1 expression in per-protocol kidney biopsies. Changes in the uAPI following transplantation were assessed using a linear mixed-effects model with time (modelled as 4-knot restricted cubic spline (B) or as a factor variable (C-E)) as fixed effect, participant as a random intercept and time as a random slope. For panel D and E an interaction term between delayed graft function or tubular VCAM1 expression was included. N=126 for panels A-D and N=26 for panel E.

#### Figure S8: The uAPI in NBCn1 WT and KO mice during acute $NH_4Cl$-loading

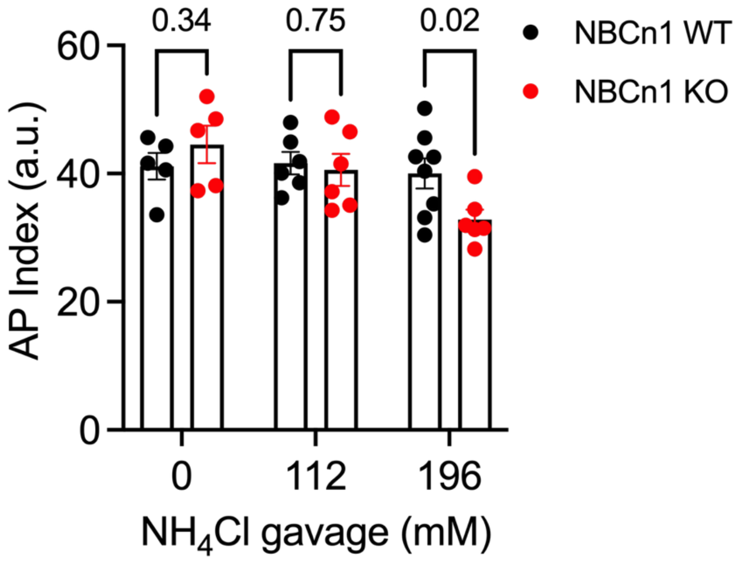

The uAPI in NBCn1 WT and KO mice after acute gavage loading of 0, 112, or 196 mM $NH_4Cl$ water (20 µl/g). Statistical differences were assessed by two-way ANOVA followed by Sidak's multiple comparisons test.

#### Figure S9: Excretion-based uAPI in NBCe-1A/B, GS, NBCn1 and RhCg+RhBg mice

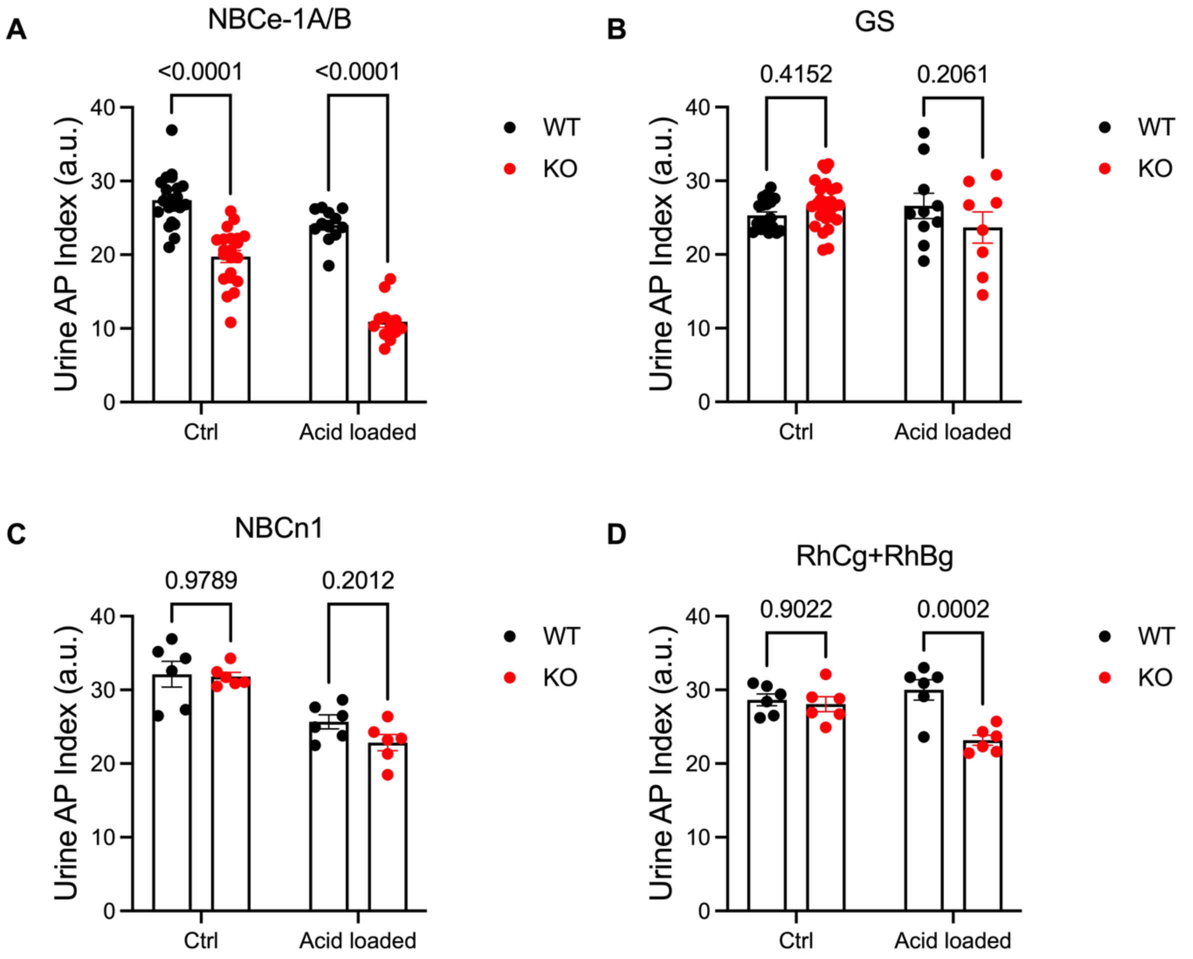

Excretion-based uAPI during control conditions and after 24 hours of acid-loading in A) NBCe-1A/B WT (n=12-21) and KO mice (n=12-21), B) GS WT (n=10-19) and KO mice (n=8-26), C) NBCn1 WT (n=6-14) and KO mice (n=6-11), and D) RhCg + RhBg WT (n=6) and KO mice (n=6). Mice were either dietary acid-loaded (0.4 mmol HCl/g chow, NBCe1, GS and RhCg + RhBg mice) or by $NH_4Cl$-enriched drinking water (196 mmol/L, NBCn1 mice). Statistical differences were assessed by two-way ANOVA followed by Sidak's multiple comparisons test.

#### Figure S10: Age-Dependent Changes in Proteinuria and the uAPI in Podocin Mice

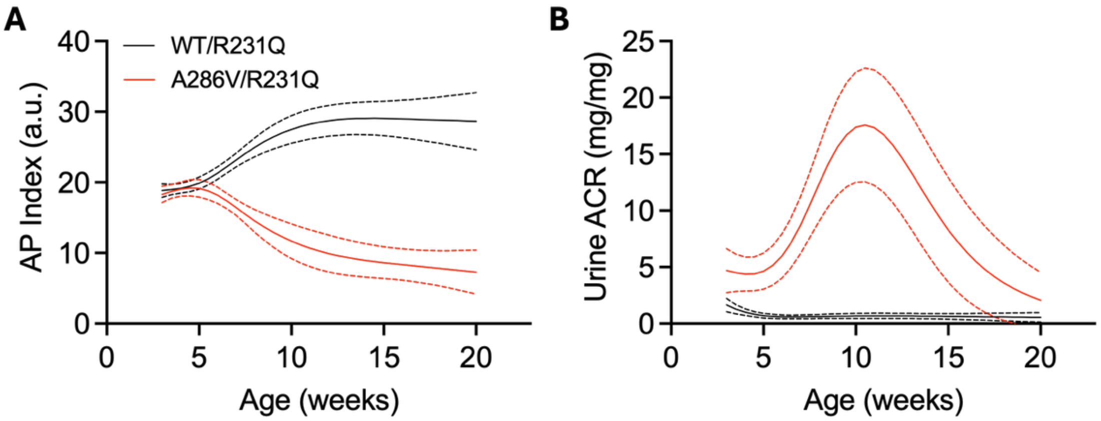

A) The uAPI and B) urine albumin-creatinine-ratio as a function of age in podocin$^{A286V/R231Q}$ (red) and WT mice (black). Changes in uAPI and uACR as a function of age were assessed using a linear mixed-effects model with age (modelled as a 4-knot restricted cubic spline), genotype and age-by-genotype as fixed effects, sex as a covariate, mouse ID as random intercept and age as random slope.

#### Figure S11: Characteristics of Doxorubicin and Vehicle-Treated Mice

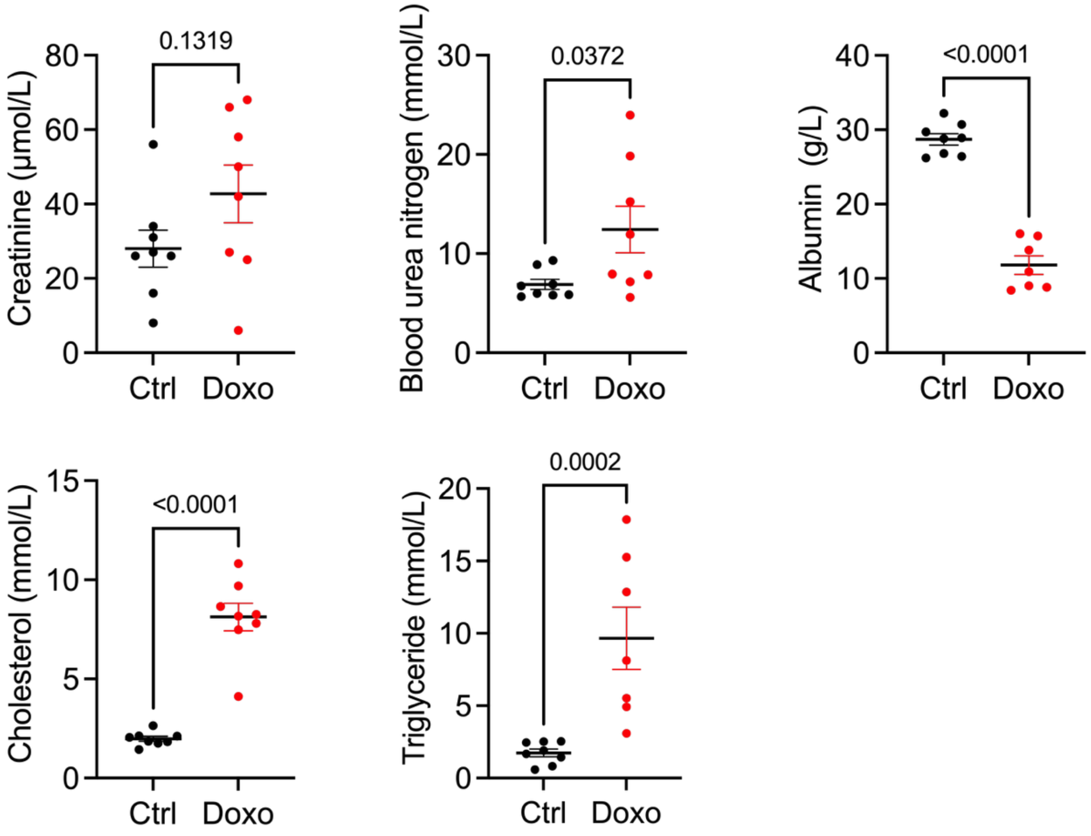

Plasma creatinine, blood urea nitrogen, albumin, cholesterol, and triglycerides in control (ctrl, black) and doxorubicin (doxo, red) treated mice measured 7 days after injection. Statistical differences were assessed using Welch’s tests. Blood urea nitrogen, cholesterol and triglycerides were log-transformed before statistical evaluation.

#### Figure S12: The effect of angiotensin II on blood pressure, albuminuria and GFR

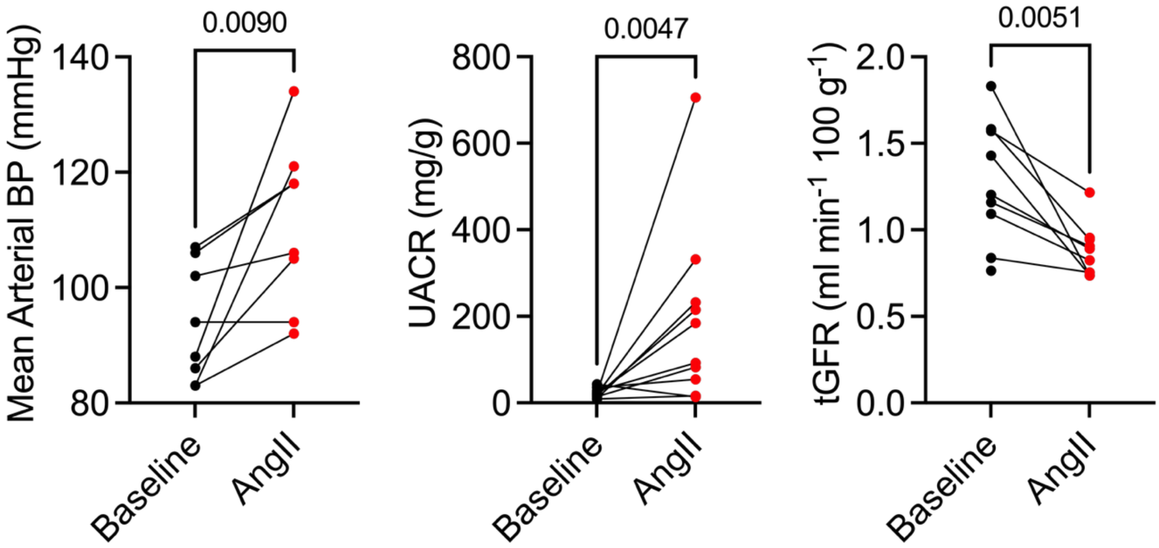

Change in mean arterial blood pressure (measured by tail-cuff), urine albumin-creatinine-ratio (uACR) and measured GFR in C57Bl/6J mice treated with continuous infusion of angiotensin II (AngII, 750 ng/kg/min) via subcutaneous osmotic mini-pumps for 4 weeks. Significance was assessed using students t-tests. UACR was log-transformed before statistical assessment.

#### Figure S13: Association between uAPI and fibronectin, glomerulosclerosis, KIM1, and measured GFR in ZSF1 rats

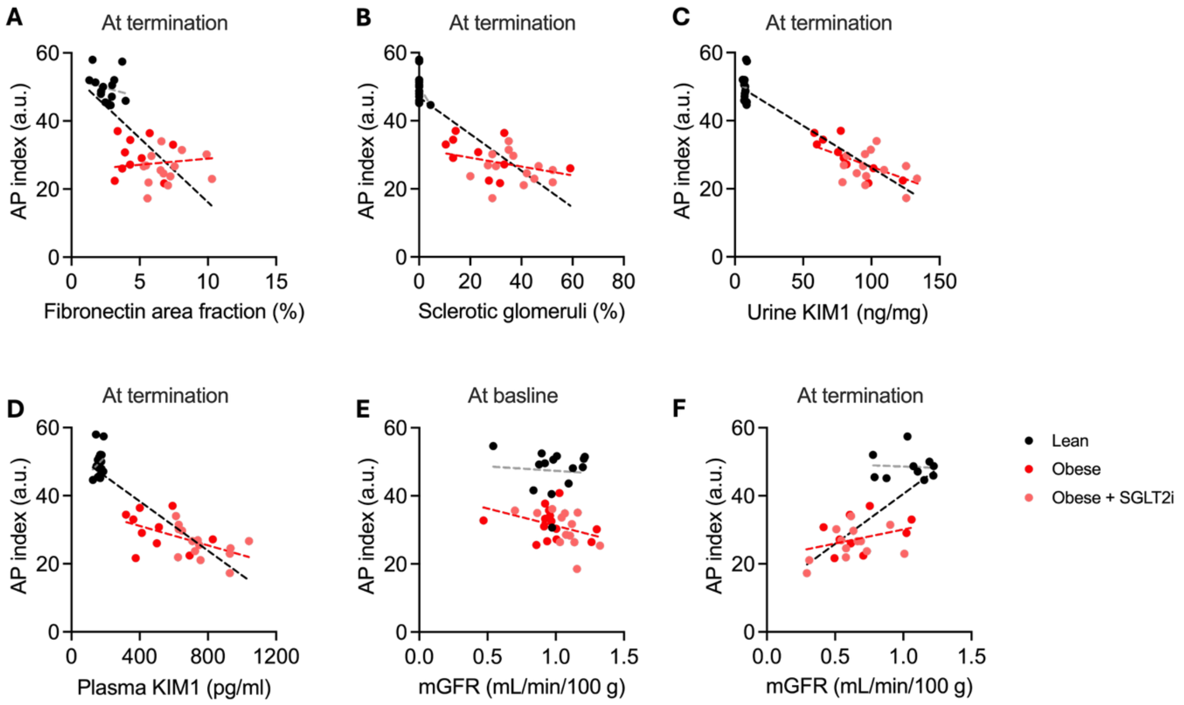

A-B) Association between A) fibronectin area and B) % sclerotic glomeruli in kidney sections and uAPI at termination (week 29-30) of obese (red), obese empagliflozin-treated (light red) and lean (black) ZSF1 rats. C-D) Association between C) urine and D) plasma KIM-1 in obese (red), obese empagliflozin-treated (light red) and lean (black) ZSF1 rats. E-F) Association between measured GFR and uAPI at E) baseline and F) termination in obese (red), obese empagliflozin-treated (light red) and lean (black) ZSF1 rats. Statistical significance was assessed using simple linear regression in lean rats and obese rats with adjustment for treatment regime (vehicle vs. empagliflozin). No significant associations were found in lean (p-values: A: 0.55, B: 0.21, C:0.99, D: 0.45, E: 0.80, F: 0.85) and only for urine and plasma KIM-1 in obese rats (p-values: A: 0.42, B: 0.69, C: 0.006, D: 0.048, E: 0.71, F: 0.11).

#### Figure S14: Association between the uAPI and additional clinical markers

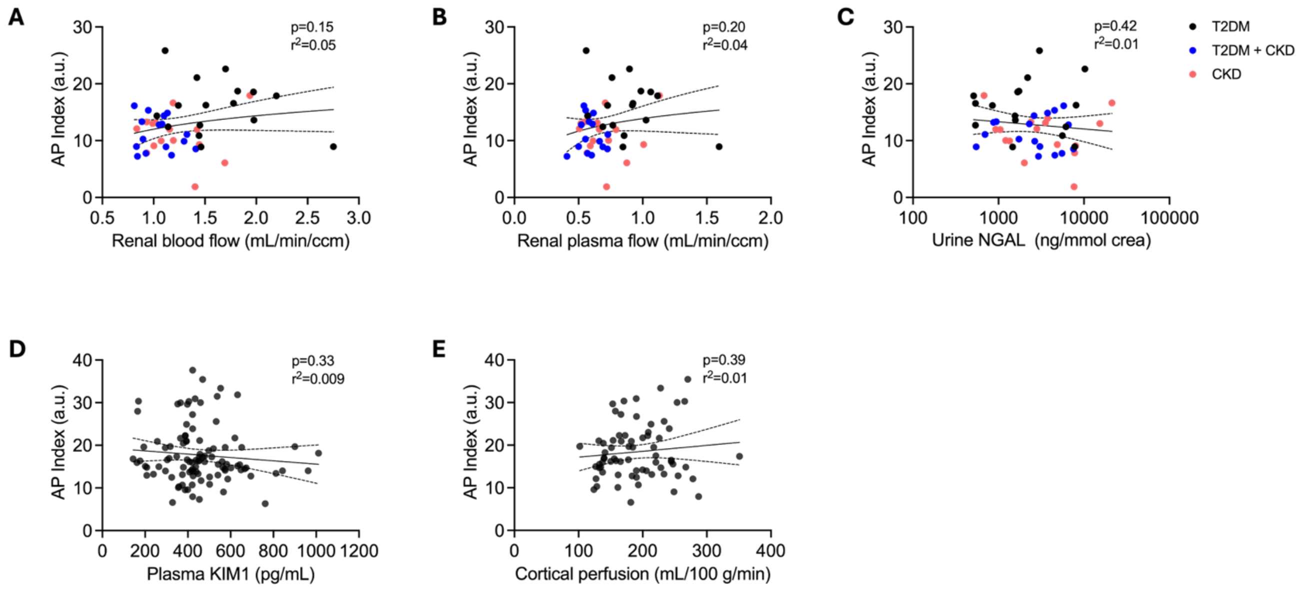

A-C) Association between the uAPI and renal blood flow (A), renal plasma flow (B) and urine NGAL (C) in the SiRENA cohort. No significant effect modifications by patient group were found. D-E) Association between plasma KIM1 (D) and cortical perfusion (E) in the SEMPA study of T2DM patients. Associations were assessed using simple linear regression. In panel A-C, possible effect modification was assessed by including an interaction term between patient type (T2DM, T2DM + CKD or CKD) and renal blood flow/renal plasma flow/urine NGAL.

#### Figure S15: Competing risk models for renal outcomes in patients with T2DM

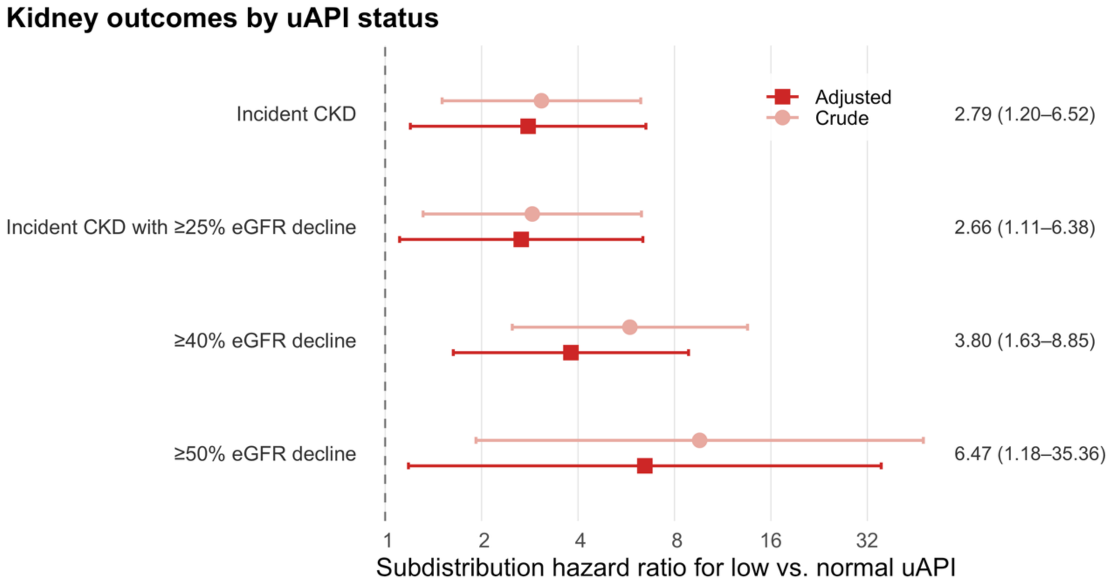

Adjusted subdistribution hazard ratios for renal outcomes in patients with type II diabetes from the SiRENA and SEMPA cohorts. Subdistribution hazard ratios were calculated from Fine-Gray competing risk models with all-cause mortality as competing risk adjusted for age, sex, BMI, diabetes status, eGFR, uACR, and systolic blood pressure.

#### Figure S16: Differential expression of $NH_4^+/NH_3$ and $H^+/HCO_3^-$ transporters and channels related to the thick ascending limb and collecting duct

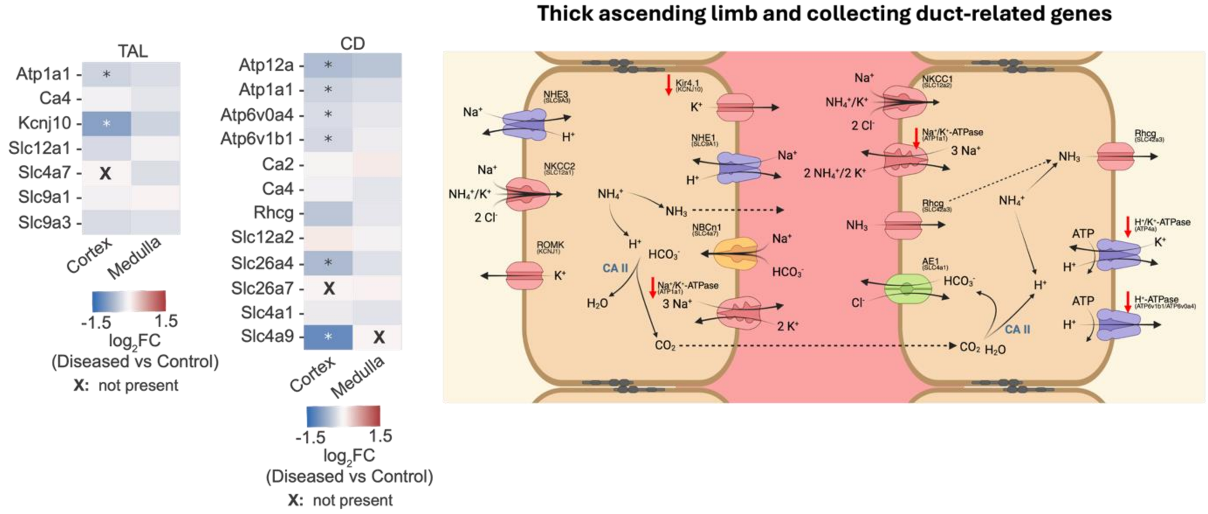

Differences in protein expression of selected proteins related to thick ascending limb or collecting duct ammonium or $H^+/HCO_3^-$ transport in cortical and medullary tissue samples from control and podocin$^{A286V/R231Q}$ mice of 10 age weeks as well as a graphical depiction of thick ascending limb and alpha-intercalated cells with red arrows arrows indicating downregulation in podocin$^{A286V/R231Q}$ mice. Proteins with a Benjamini-Hochberg false discovery rate <0.05 (Welch's t-tests, corrected within the pre-specified protein set) and |$\log_2$ fold change| >0.2 are annotated with an asterisk and arrows in the schematic. X: not found.

#### Figure S17: Differential expression of genes related to $NH_4^+/NH_3$ or $H^+/HCO_3^-$ transport in proximal tubule cells of control and patients with type II diabetes

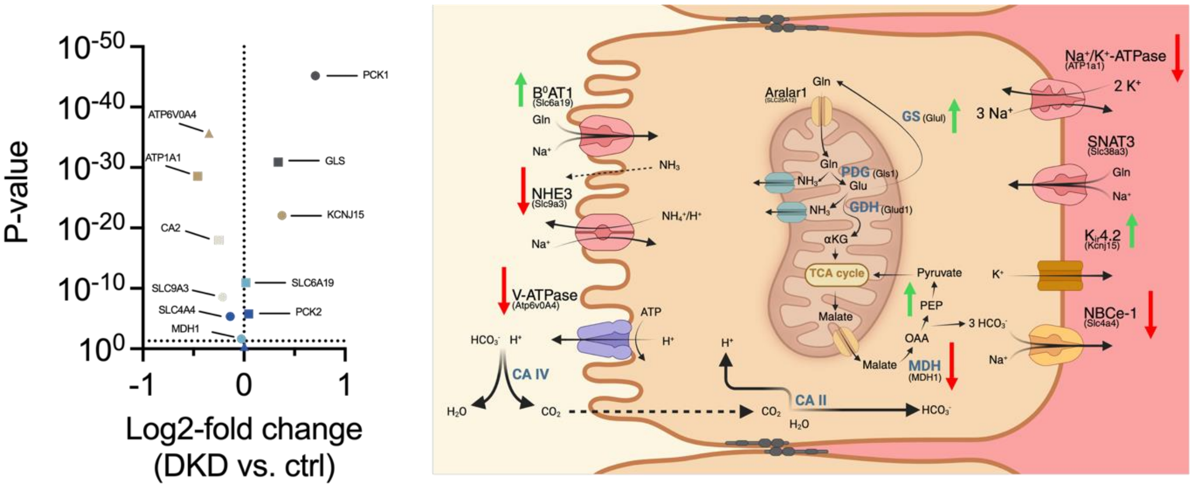

Differences in proximal tubule cell mRNA of selected genes related to ammoniagenesis or ammonium or $H^+/HCO_3^-$ transport in patients with diabetes vs. controls as well as graphical depiction of a proximal tubule cell with arrows indicating up-regulation (green) or down-regulation (red) in the diabetes patients. Data was extracted from a publicly available dataset of single cell sequencing of human kidney samples[6].

#### Figure S18: Differential expression of genes related to $NH_4^+/NH_3$ or $H^+/HCO_3^-$ transport in thick ascending limb and type A-intercalated cells in controls and patients with type II diabetes

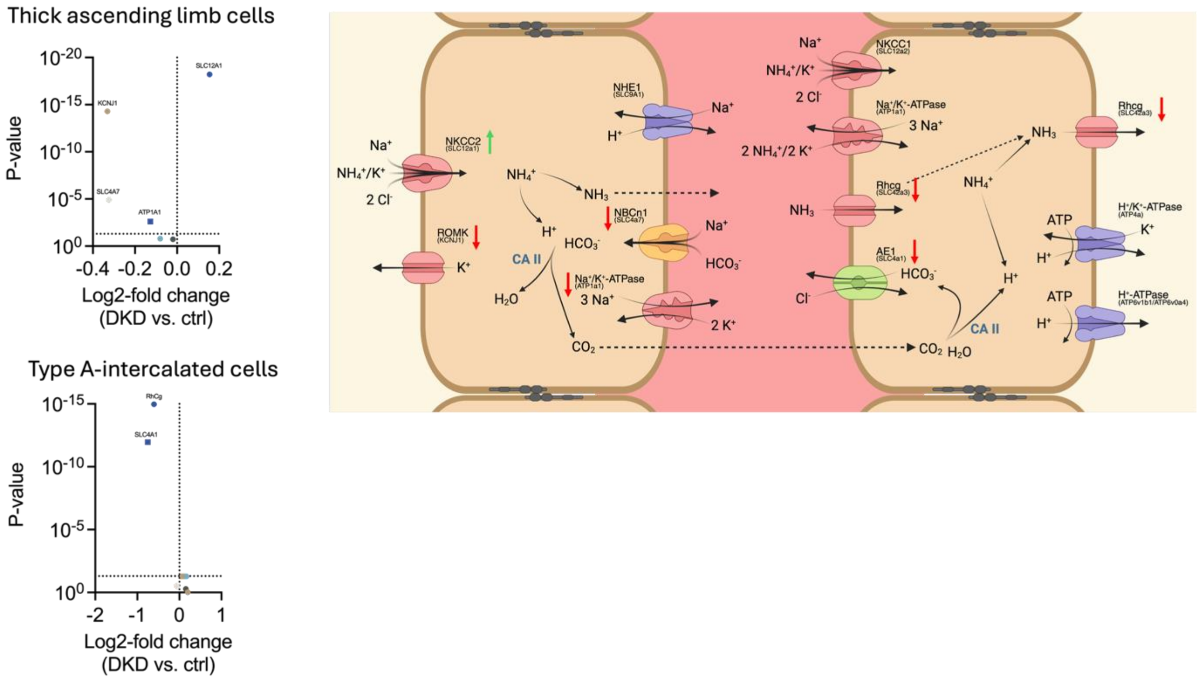

Differences in thick ascending limb and alpha-intercalated cell mRNA of selected genes related to ammonium or $H^+/HCO_3^-$ transport in patients with diabetes vs. controls as well as graphical depiction of thick ascending limb and alpha-intercalated cells with arrows indicating up-regulation (green) or down-regulation (red) in the diabetes patients. Data was extracted from a publicly available dataset of single cell sequencing of human kidney samples[6].

#### Figure S19: Differential expression of genes related to $NH_4^+/NH_3$ or $H^+/HCO_3^-$ transport in injured proximal tubule cells

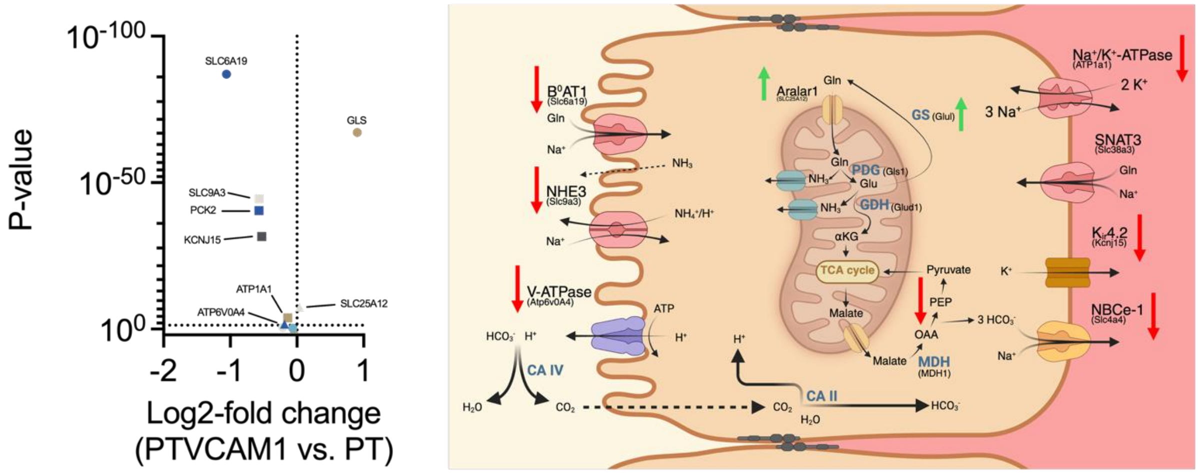

Differences in proximal tubule cell mRNA of selected genes related to ammoniagenesis or ammonium or $H^+/HCO_3^-$ transport in injured proximal tubule cells vs. normal proximal tubule cells as well as graphical depiction of a proximal tubule cell with arrows indicating up-regulation (green) or down-regulation (red) in injured proximal tubule cells. It was not possible to plot SLC4A4 in the volcano plot because the published p-value was "0". SLC4A4 had a log2-fold change of -2.03. PCK1 is not plotted in the volcano plot but had a p-value 6.282e-182 and a log2-fold change of -1.6. Data was extracted from a publicly available dataset of single cell sequencing of human kidney samples[6].

#### Figure S20: Subgroup analyses for low vs. normal uAPI across RENVAS, PUMA and SLEEP

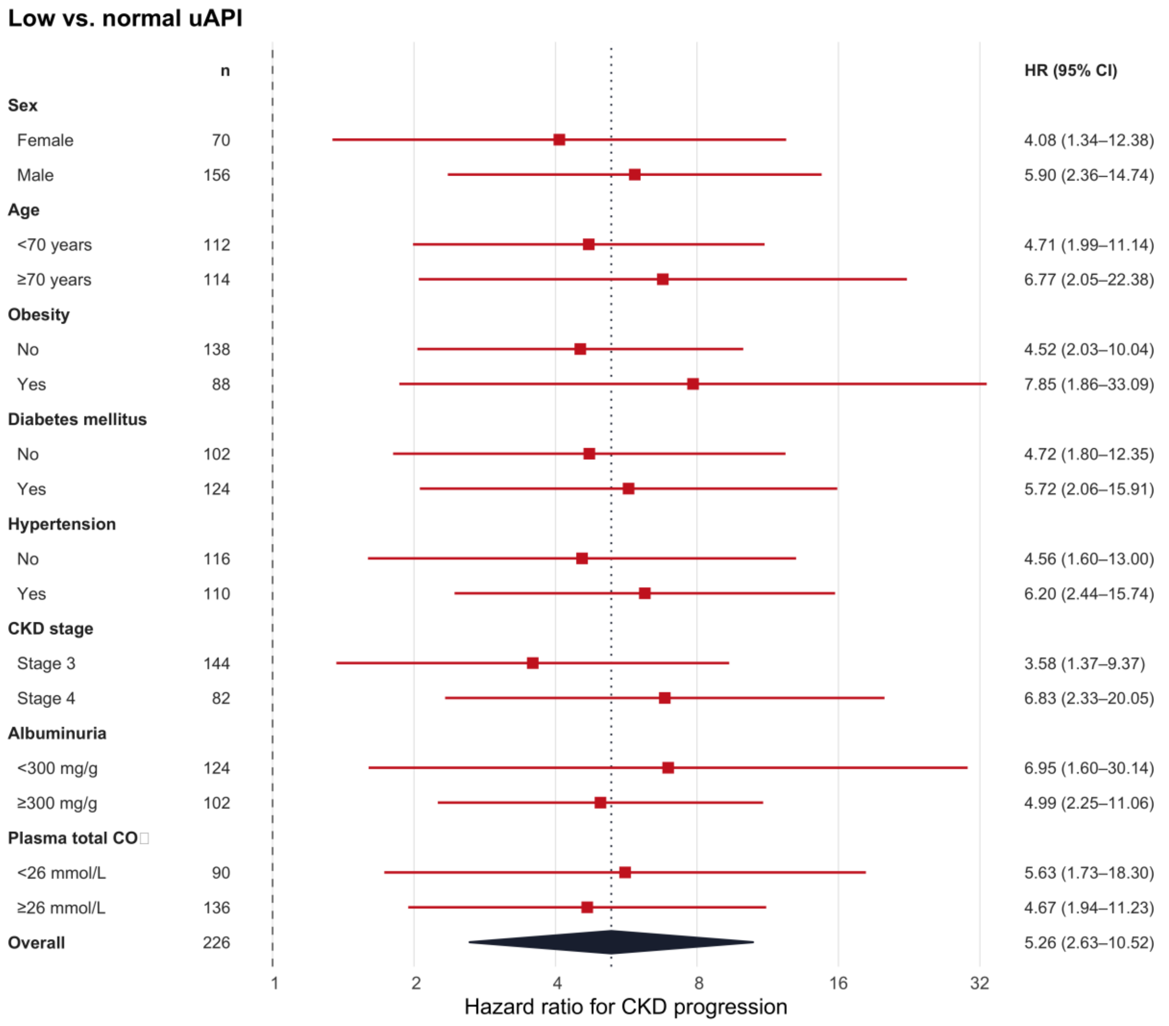

Unadjusted hazard ratios for CKD progression in RENVAS, PUMA and SLEEP for low vs. normal uAPI. Hazard ratios were estimated from unadjusted cox proportional hazards stratified by cohort.

#### Figure S21: Subgroup analyses per SD lower uAPI across RENVAS, PUMA and SLEEP

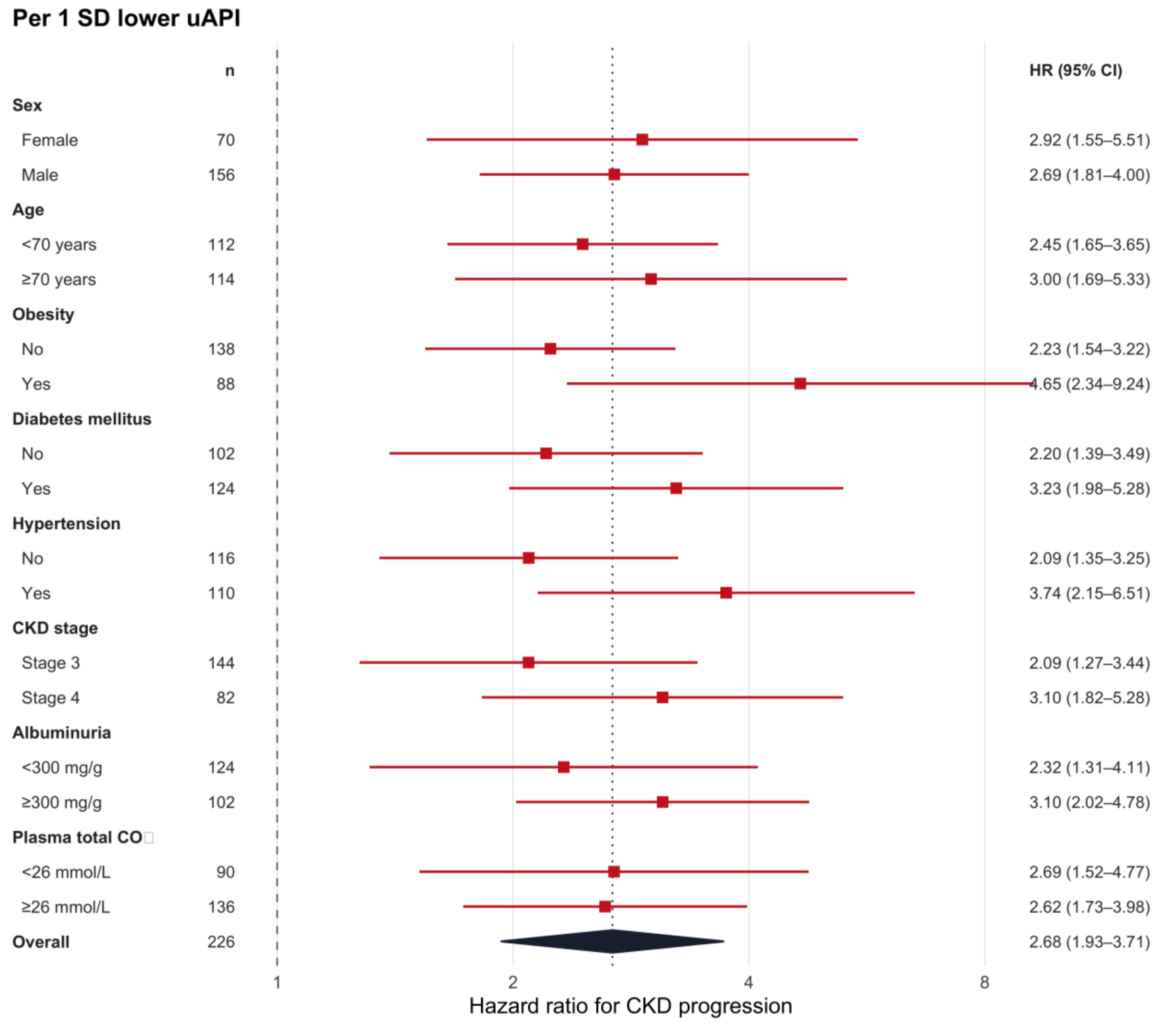

Unadjusted hazard ratios for CKD progression in RENVAS, PUMA and SLEEP per SD lower uAPI. Hazard ratios were estimated from unadjusted cox proportional hazards stratified by cohort.

#### Figure S22: Competing risk models for renal outcomes and CKD progression in CKD

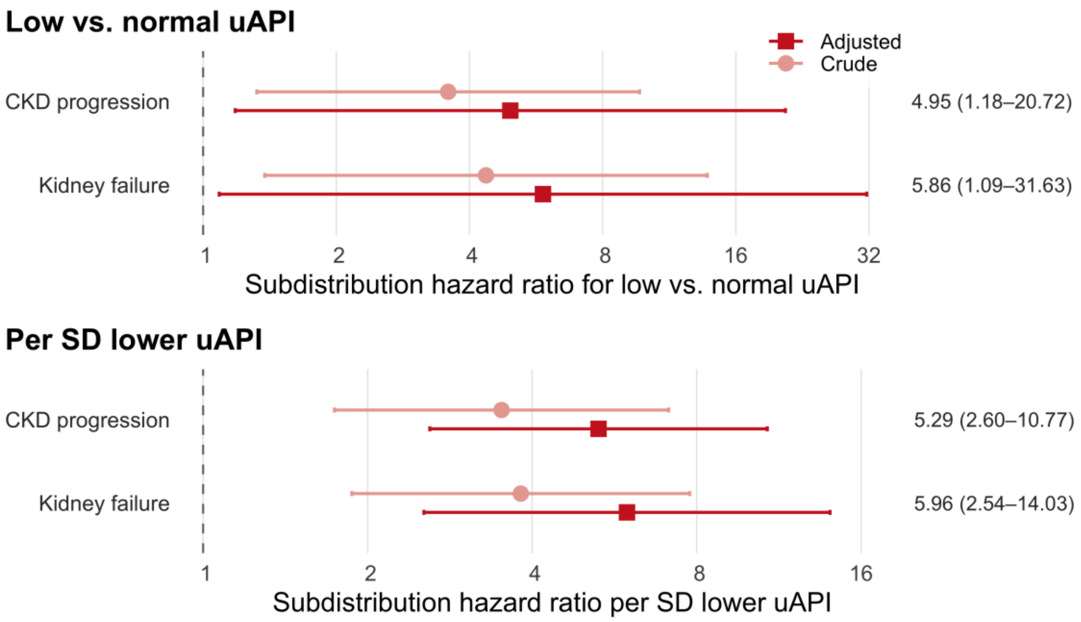

Crude and adjusted subdistribution hazard ratios for renal outcomes in CKD patients from the SLEEP cohort (n=75). Subdistribution hazard ratios were calculated from Fine-Gray competing risk models with all-cause mortality as competing risk adjusted for age, sex, BMI, eGFR, uACR, systolic blood pressure, and total $CO_2$.

#### Figure S23: Calibration of the KFRE and KFRE + uAPI in RENVAS + PUMA and SLEEP

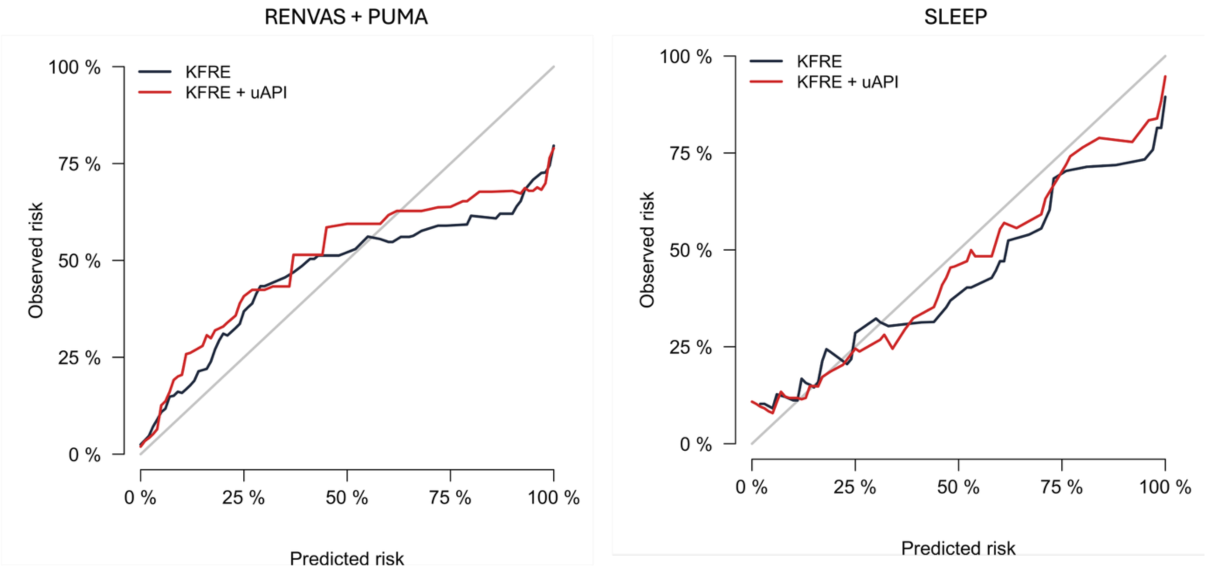

Calibration curve for RENVAS + PUMA and SLEEP of the KFRE and KFRE + uAPI.

In RENVAS + PUMA, calibration slopes at year 5 were 0.771 (95% CI: 0.57 to 0.97, KFRE) and 0.769 (95% CI: 0.58 to 0.96, KFRE + uAPI), observed risk was 29.9% and predicted risks were 28.9% (KFRE, O/E: 1.03) and 27.5% (KFRE + uAPI, O/E: 1.09).

In SLEEP calibration slopes at year 4 were 1.14 (95% CI: 0.77 to 1.5, KFRE) and 1.19 (95% CI: 0.84 to 1.54, KFRE + uAPI), observed risk was 41.8% and predicted risks were 46.1% (KFRE, O/E: 0.906) and 44% (KFRE + uAPI, O/E: 0.95).

#### Figure S24: Discrimination and reclassification metrics in RENVAS + PUMA and SLEEP

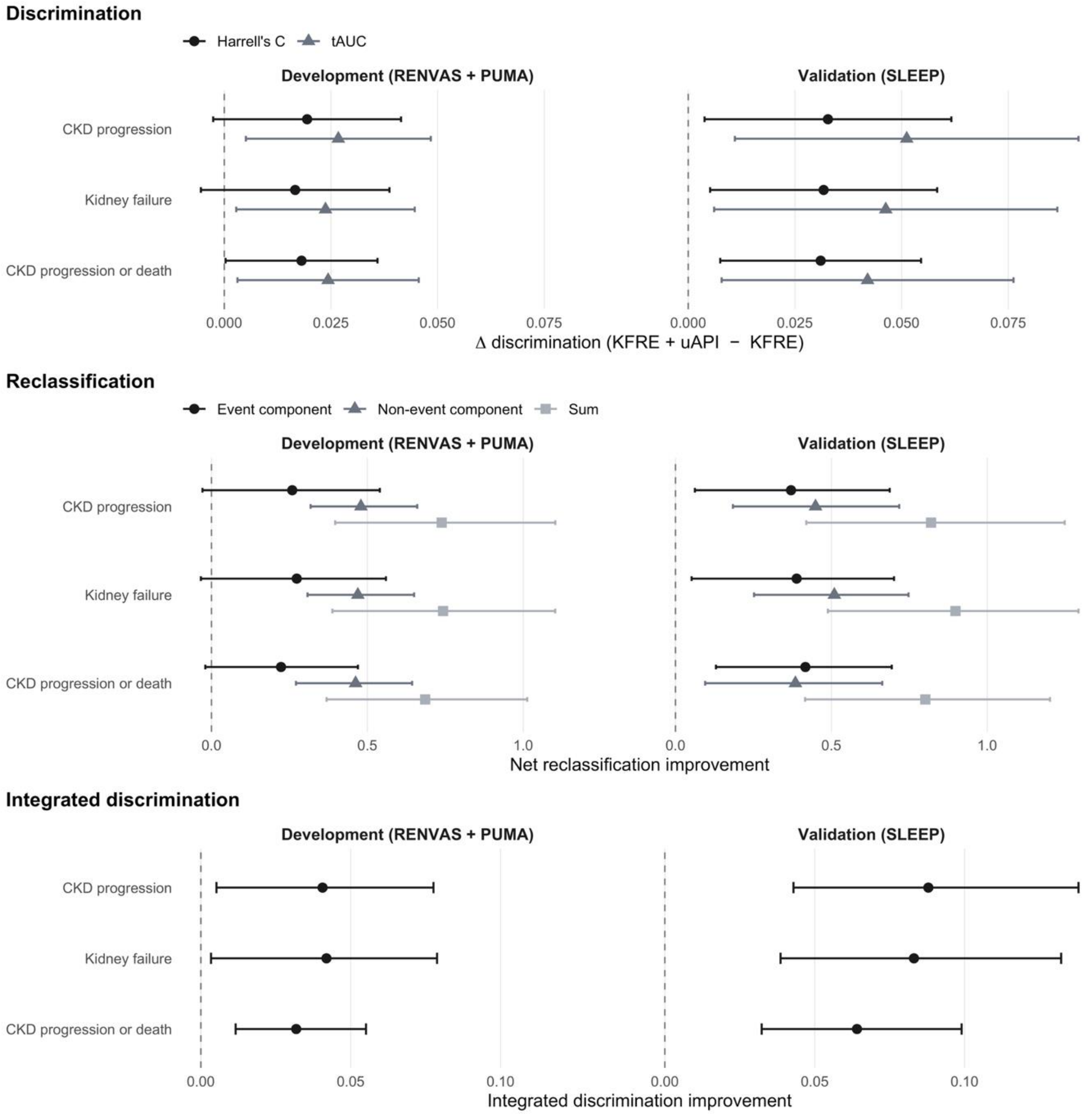

Difference between the two models in discrimination (Harrell's C and time-dependent AUC), net reclassification improvement (event component, non-event component, and their sum), and integrated discrimination improvement, in RENVAS + PUMA and SLEEP. The KFRE linear predictor was entered as a fixed offset, and the uAPI coefficient was estimated in RENVAS and PUMA and applied unchanged to SLEEP.

#### Figure S25: Relationship between KFRE-predicted risk, the uAPI, and treatment efficiency.

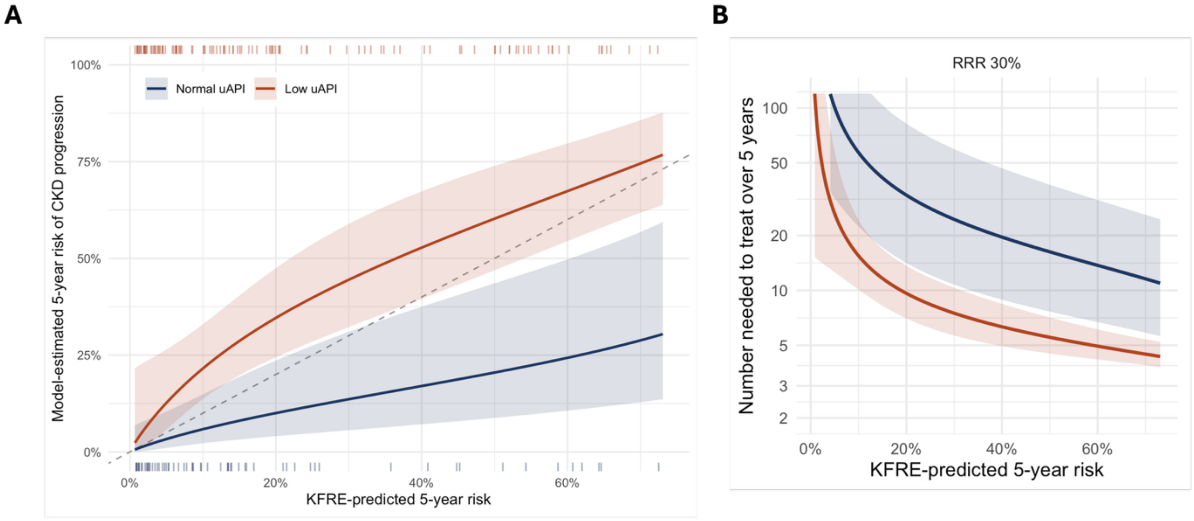

C

| | Number-needed-to-treat | | |
|---|---|---|---|
| 5-year KFRE risk | Normal uAPI | Low uAPI | Ratio |
| 5% | 101 | 32 | 3.8 |
| 10% | 57 | 22.4 | 3.7 |
| 20% | 33.1 | 14.1 | 3.5 |
| 30% | 24.4 | 10.7 | 3.3 |
| 50% | 16.2 | 7.6 | 2.9 |

(A) Model-estimated five-year risk of CKD progression as a function of KFRE-predicted five-year risk, in participants with a low (<13.4 a.u.) versus normal uAPI, pooled across RENVAS, PUMA and SLEEP (n=226). Estimated by pseudo-observation regression on the Aalen-Johansen probability with a complementary log-log link, with the two curves constrained to be parallel on that scale (an interaction model was not supported, Wald p=0.23). Shaded areas are 95% confidence intervals, the dashed line indicates agreement with the KFRE, and ticks show the distribution of KFRE-predicted risk (normal uAPI below the axis, low uAPI above). Curves are shown only over the range of predicted risk covered by both strata. (B) Number needed to treat over five years to prevent one CKD progression event, derived from the curves in (A) assuming a constant 30% relative risk reduction. Curves truncated at 120. (C) Number needed to treat at selected values of KFRE-predicted risk, with the ratio between uAPI strata.The parallel constraint makes the two curves converge as predicted risk approaches zero, the tertile analysis (Table S8) shows separation in the lowest tertile.
